# Systematic review: Longitudinal effects of the COVID-19 pandemic on child and adolescent mental health

**DOI:** 10.1101/2022.12.20.22283720

**Authors:** Kristin Wolf, Julian Schmitz

## Abstract

**Background:** The COVID-19 pandemic and the protection measures to contain its spread have massively changed daily lives of billions of children and adolescents worldwide.

**Objective:** We conducted a systematic review to investigate the global longitudinal effects of the COVID-19 pandemic on various mental health outcomes in children and adolescents over a period of one and a half years.

**Methods:** This review was conducted in accordance with the guidelines recommended by the Preferred Reporting Items for Systematic Reviews and Meta-Analyses (PRISMA) statement. The databases “PubMed”, “Web of Science”, and “APA PsycInfo” were searched (last access: 01/2022). Studies were included if they were peer-reviewed and published between December 2019 and December 2021, if they followed a longitudinal or repeated cross-sectional design, and if they assessed the effect of the COVID-19 pandemic or a related stressor on mental health indicators.

**Findings:** Of 7,451 identified studies, 69 studies (*n* ∼ 130,000) meeting eligibility criteria were included in a qualitative analysis. The results indicate a general trend of decreased psychological well-being, increased psychopathological distress, and heightened symptom levels (particularly of depressive and anxiety symptoms) from before to during the pandemic. Data suggests that both the intensity of protection measures and infection dynamic were positively associated with severity of psychopathology. The most reported influencing factors on the effect of the pandemic on child and adolescent mental health were age, gender, socio-economic status, previous state of mental and physical health, self-regulation abilities, parental mental health, parenting quality, family functioning, social support, isolation and loneliness, health-related worries, and consistent routines and structure.

**Conclusion:** Our systematic review shows that children and adolescents worldwide have experienced increased psychological distress due to the COVID-19 pandemic. These results call for improvement in access to child and adolescent mental health care and the prioritisation of child and adolescent well-being in political decision making.

## Introduction

### The COVID-19 pandemic

In December 2019, the outbreak of a respiratory disease caused by the novel virus “Severe Acute Respiratory Syndrome Coronavirus type 2” (SARS-CoV-2) was reported in the People’s Republic of China [1]. The outbreak rapidly developed into an epidemic in the country by January 2020. The virus quickly spread to other regions of the world. On 11 March 2020, The World Health Organization (WHO) [1] declared the spread of the virus-induced disease “COVID-19” as a global pandemic. By October 2022, more than 610 million confirmed cases of COVID-19 were reported to the WHO, of which over 6.5 million were fatal. As a result of the rapid spread of infection, drastic measures were taken worldwide to contain the spread of the virus, leading to massive restrictions on everyday life and the global economic crisis of 2020/21.

### Effects of the COVID-19 pandemic on the daily life of children and adolescents

For most children and adolescents, the current pandemic represents a first confrontation with severe threat of disease, and potential death and grief in their lives [2–4]. In many the pandemic has caused anxiety and worries about infection and the health of themselves and of their family and friends [3,5]. Feelings of safety, trust, and security have been replaced by a perception of the world as a dangerous and unsafe scary place [2,6,7]. According to a British survey conducted by Youngminds (2021) [7], young people reported loneliness and isolation, concerns about school work, and breakdown of routines to be the most stressful experiences in the pandemic.

The global economic crisis caused by pandemic-related restrictions has led to a sharp rise in unemployment and poverty around the world [8,9]. Especially in low-income countries, these economic hardships forced many children into exploitative and dangerous work to support their families [8].

Further, since the start of the pandemic, essential health services have been disrupted in 90% of countries worldwide [10]. This means that children’s and adolescents’ access to routine immunizations and examinations has been impaired, leading to increased morbidity and mortality [9,10]. Many psychiatric and psychosomatic in- and outpatient clinics were partially or fully closed for multiple weeks. The work of child protection services has been impaired in at least 104 countries, making it hard to prevent, report, detect, and respond appropriately to cases of child maltreatment [11,12].

One of the biggest and most far-reaching changes to the lives of children and adolescents in the pandemic has been the closure of schools and childcare facilities to prevent the spread of the virus. These closures have been globally regarded as necessary due to the impracticability of distancing practices in school because of limited space, frequent and varied interactions among large groups of pupils, and difficulties for (especially younger) children to follow hygiene and distancing guidelines [13]. According to the World Health Organization (2022) [10], the closures of schools have led to the largest disruption of education systems in history, affecting nearly 1.6 billion students in more than 190 countries.

Regarding the closer social environment of children and adolescents, it is likely that social relationships of children and adolescents have suffered during the pandemic due to limited and discouraged socialising and imposed isolation from friends and extended family members [11,14].

Not only children and adolescents but entire family systems have experienced disrupted daily activities and pandemic-related stress. Therefore, inner-familiar tension has increased and family dynamics in many households have changed [6,9].

Taken together, the COVID-19 pandemic and related containment measures have massively changed the daily lives of children and adolescents. For many, normal development has been impaired, and stress and strain have increased while the availability of many coping, support, and protection resources has been limited.

### Research on the effect of the COVID-19 pandemic on child and adolescent mental health

Since the beginning of the COVID-19 pandemic in spring 2020 a lot of research investigating its mental health consequences has been conducted. Some reviews and meta-analyses have already tried to summarise this enormous body of research.

Santomauro et al. (2021) [15] estimated that there has been an increase of 27.6% in depression and of 25.6% in anxiety disorders in the general population due to the pandemic. In addition, a number of reviews and meta-analyses which assessed the psychological consequences of the COVID-19 pandemic on different age groups have found a strong increase of mental health problems in children and adolescents [6,16–22].

However, it must be taken into consideration that previous reviews and meta-analyses included mostly cross-sectional studies which makes it difficult to distinguish between pre-existing levels of psychopathology and the pandemic-specific consequences and can lead to an overestimation of effects.

Therefore, longitudinal studies seem more appropriate for investigating these effects, as they allow a direct detection of developmental pathways across time. Unfortunately, the few meta-analyses which use longitudinal assessments of mental health symptoms focus on the general population and not on children and adolescents in particular [23–25].

### The current review

The current study attempts to fill in these gaps in research by reviewing only longitudinal and repeated cross-sectional studies on the mental health consequences of the COVID-19 pandemic on child and adolescent mental health published over a course of two years. The objectives are to isolate the pandemic’s effects on a broad spectrum of mental health outcomes, to learn about factors influencing these effects, to investigate long-term psychological consequences over the course of the pandemic, and to create a large sample of studies conducted in many different countries to obtain a higher generalizability of findings. This study might also be beneficial to the disaster theory and research as the COVID-19 pandemic significantly differs from disasters frequently studied until now regarding duration, intensity, and differential effects.

The following research questions are investigated in this paper:

1) How did the frequency of psychological distress in the general population of children and adolescents change from before the COVID-19 pandemic to during the pandemic?
2) How did the frequency of psychological distress in the general population of children and adolescents develop over the course of the pandemic?
3) What are moderating and mediating factors that influence the potential effects of the COVID-19 pandemic on child and adolescent mental health?

Psychological distress serves as an umbrella term comprising both global measures of mental health and psychopathology (i.e., quality of life, well-being, life satisfaction, psychological stress, and affect) and symptoms of common mental health disorders in childhood and adolescence. These involve internalising symptoms (including depression and anxiety), externalising symptoms (including hyperactivity, inattention, and disruptive beahviour problems), psychosomatic complaints, addictive behaviour (including substance abuse and excessive use of electronic media), and post-traumatic stress symptoms.

These symptom categories match commonly described outcomes in research discussed previously and cover a broad range of mental health problems which can occur in childhood and adolescence.

In the view of the literature discussed above the following assumptions were derived regarding the three research questions:

1) Exposure to the COVID-19 pandemic is expected to result in an increase of general psychopathological stress levels and symptoms of mental disorders in children and adolescents compared to pre-pandemic levels, particularly internalising symptoms of depression and anxiety, externalising symptoms, attentional problems, post-traumatic stress, and psychosomatic symptoms.
2) Pandemic-related protection measures, particularly confinement and quarantine, are expected to have a negative effect on children‘s and adolescents‘ mental health. Reducing restrictions over the course of the pandemic should therefore be associated with decreasing symptoms of poor mental health.
3) Disadvantaged children and adolescents are expected to be more severely affected by the psychological effects of the COVID-19 pandemic. This includes children and adolescents who are dependent on special support due to precarious family, economic and social circumstances, disability, or illness.

## Methods

### Literature research

This review was carried out in accordance with the guidelines proposed by the Preferred Reporting Items for Systematic Reviews and Meta-Analyses (PRISMA) statement [26]. There was no registered protocol.

The literature research was conducted within the literature databases “Web of Science”, “PubMed”, and “APA PsycInfo”. Each of these sources were consulted the last time on 12 January 2022. The following search strategy was used to identify studies examining the effect of the COVID-19 pandemic on child and adolescent mental health: “(COVID-19 OR corona* OR SARS-CoV-2 OR novel coronavirus) AND (psychological OR psychopatholog* OR mental health OR psychiatric disorders OR interna* OR externa* OR depress* OR anxi* OR sleep* OR hyperact* OR ADHD OR attention OR autis* OR neurodevelopment* OR disruptiv* OR conduct OR defiant OR oppositional OR emotional OR behavio* OR addict*) AND (adolescent* OR child* OR youth OR minor* OR teenager* OR juvenile*)”. To ensure sufficient data, a broad range of mental health symptoms were initially included in the search terms.

If available at the respective databases, the following automatic filters were used: The search terms named above had to appear in the title and/or abstract of the studies, studies had to be peer-reviewed journal articles written in the English language and published between December 2019 and December 2021, participants had to be aged between 0 and 18 years, and methodology was restricted to quantitative designs.

Additional studies were identified via the ancestry approach. For this purpose, reference lists of studies included in the systematic review and of reviews and meta-analyses on the same topic were examined.

The identified studies were then manually screened multiple times by one author based on titles, abstracts, and methodology (i.e., longitudinal, repeated, or one-time cross-sectional). The remaining studies were then textually reviewed, and results were summarised in a table.

To narrow down the number of findings further, it was decided to only include studies with a longitudinal or repeated cross-sectional research design with at least one measurement during the pandemic and only studies which used community samples.

The process of study selection is depicted in Fig 1.

**Fig 1.**
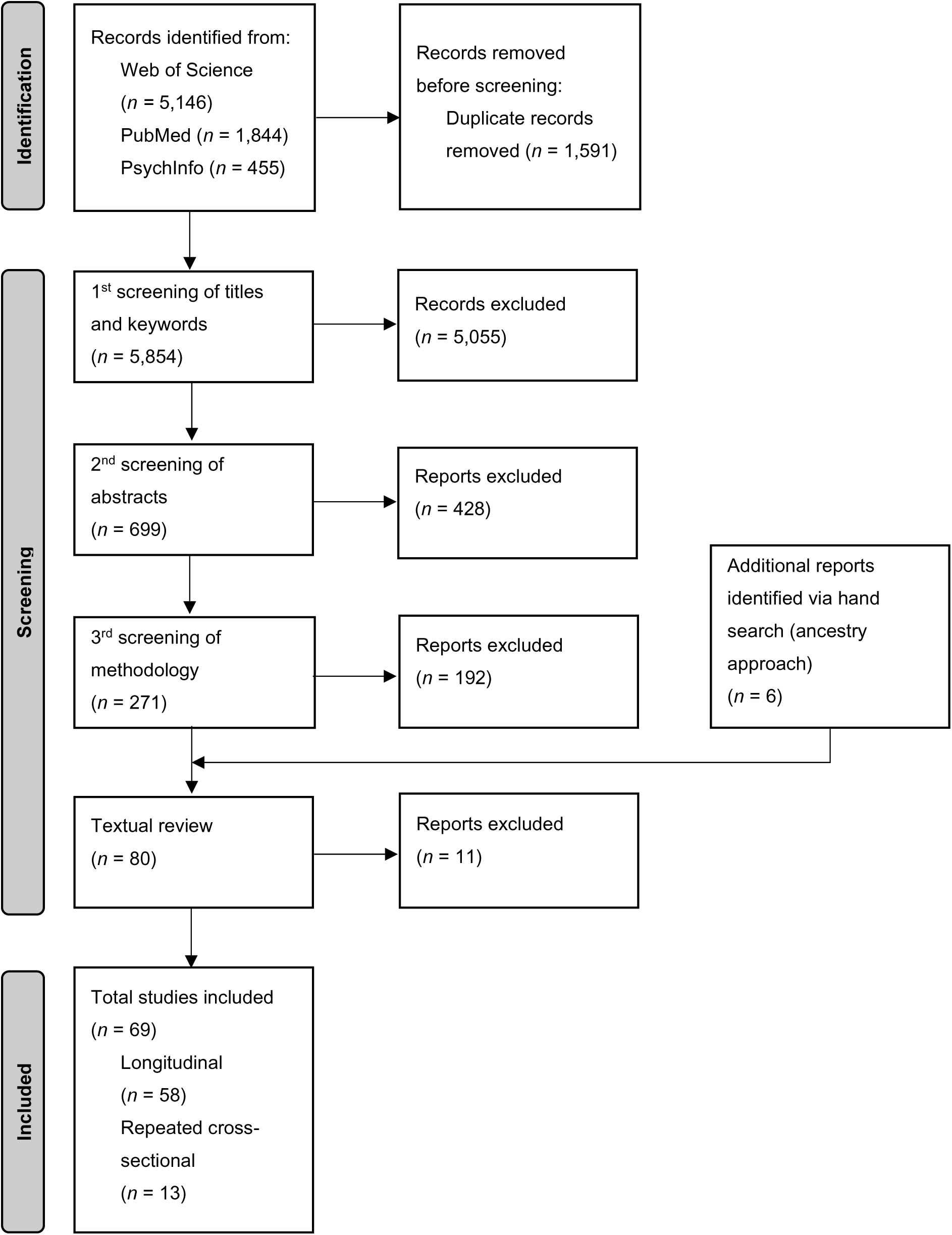
Process and Results of Literature Research. Adapted from Page et al. (2021) [26]

### Synthesis of results

To synthesise the body of research supporting changes in distinct mental health outcomes more explicitly, the findings of studies comparing measures before and during the pandemic were sorted by examined outcome (e.g., depressive symptoms) and reported change in the outcome (i.e., increase, no change, or decrease from before to during the pandemic). The number of studies examining each mental health indicator and the number of findings for each possible direction of change were calculated and tabulated.

If possible, outcomes measuring overlapping constructs and symptom categories were grouped together to create larger pools of studies investigating each indicator to allow for more profound conclusions. In detail, the following outcomes were summarised as one mental health indicator, respectively:

• “Psychological well-being” and “quality of life” were grouped together.
• Total scores of scales assessing multiple mental health outcomes were summarised as “global measures of mental health problems”.
• “Internalising symptoms/problems” and “emotional symptoms/problems” were summarised as “internalising symptoms”.
• “Externalising symptoms/problems” and “behavioural symptoms/problems” were summarised as “externalising symptoms”.
• “Conduct”, “oppositional”, “challenging”, and “impulsive behaviour symptoms” were summarised as “disruptive behaviour symptoms”.
• “Inattention”, “attentional problems”, and “sluggish cognitive tempo” were summarised as “cognitive symptoms”.

The following psychopathological indicators and symptoms were included later in addition to the outcomes chosen a priori and named above, as they appeared in studies chosen for this review: Resilience, suicidal symptoms, neurotic symptoms, dissociative symptoms, eating disorder symptoms, and psychotic symptoms.

For the examination of overall directions of changes in mental health, the number of findings supporting an increase, no change, or decrease in a mental health indicator were separately added together. For this purpose, outcomes were grouped as “positive mental health indicators” if they are positively associated with mental health (e.g., well-being, quality of life, life satisfaction) and as “negative mental health indicators” if they are negatively associated with mental health (e.g., stress, negative affect, psychopathological symptoms).

The results of studies investigating changes over the course of the pandemic were synthesised separately. The emphasis was thereby put on findings on the effects of certain health protection measures (i.e., lockdown, confinement) and the general severity of restrictions on mental health outcomes.

To examine the data on different influencing factors (i.e., moderators, mediators, correlates) on the effect of the pandemic on mental health, factors reported by the identified studies were collected as “COVID-19 related stressors” or “other variables” and then tabularly sorted in the following categories:

• sociodemographic factors,
• intra-individual factors,
• parental/family factors,
• social factors,
• COVID-19-related factors,
• and behavioural factors.

Two additional approaches to review the studies’ findings were used post-hoc to identify sources of variance in outcomes between studies. To find evidence on systematic effects of a) the age of participants and b) the region where the respective studies were conducted, all studies examining changes from prior to during the pandemic were sorted by a) age of the participants (< six years, six–12 years, > 12 years) and b) country, respectively. For the evaluation of age effects, findings of an increase, no change, or decrease in each mental health indicator were counted separately. For the evaluation of the influence of region, findings were only sorted by indicated direction of change in mental health and not by specific indicators due to the small number of studies for each country.

## Results

### Sample characteristics

In total, 69 studies, published between September 2020 and December 2021 and assessing around 130,000 participants, were included in this review. Fifty-eight of these studies used a longitudinal, within-subjects design. Thirteen studies applied a repeated cross-sectional, between-subjects design (e.g., cohort designs, matched convenience samples). Two of these studies analysed both within-subjects (i.e. intra-individual development) and between-subjects comparisons (i.e. age group comparisons) [27,28].

All included studies conducted their in-pandemic assessment between February 2020 and June 2021, covering a period of 17 months. The majority of studies were carried out in the months of April (*n* = 33), May (*n* = 37) and June 2020 (*n* = 29), which correlates with the first wave of the pandemic and the first lockdown in many countries. Far fewer studies were conducted after the summer of 2020 and in the first half of 2021. In 45 of the 69 included studies, data was collected during lockdown, confinement, or stay-at-home orders. Fourteen studies assessed children’s and adolescents’ mental health indicators and/or symptoms multiple times during the pandemic [27–40].

In total, data from 21 countries, comprising 14 European, four Asian, two North American countries, and one South American country, was included in our review. Most studies were conducted in the United States of America (*n* = 14), China (*n* = 8), the United Kingdom (*n* = 7), Germany (*n* = 6), and Canada (*n* = 6).

Concerning the age of the examined children and adolescents, all age groups between three and 18 years are included in this review. Hereby, early teenage years (i.e., 12 to 14 years) were most frequently included in the study population, while research investigating effects on early childhood was rather rare. More concretely, nine studies included participants aged six years or younger, 34 studies included participants aged six to 12 years, and 44 studies included participants aged above 12 years. Note that some studies fell into multiple of these three groups.

### Overview of included studies

Table 1 displays the longitudinal studies and the two studies using both longitudinal and repeated cross-sectional approaches. Table 2 depicts the remaining 11 repeated cross- sectional studies.

**Table 1.**
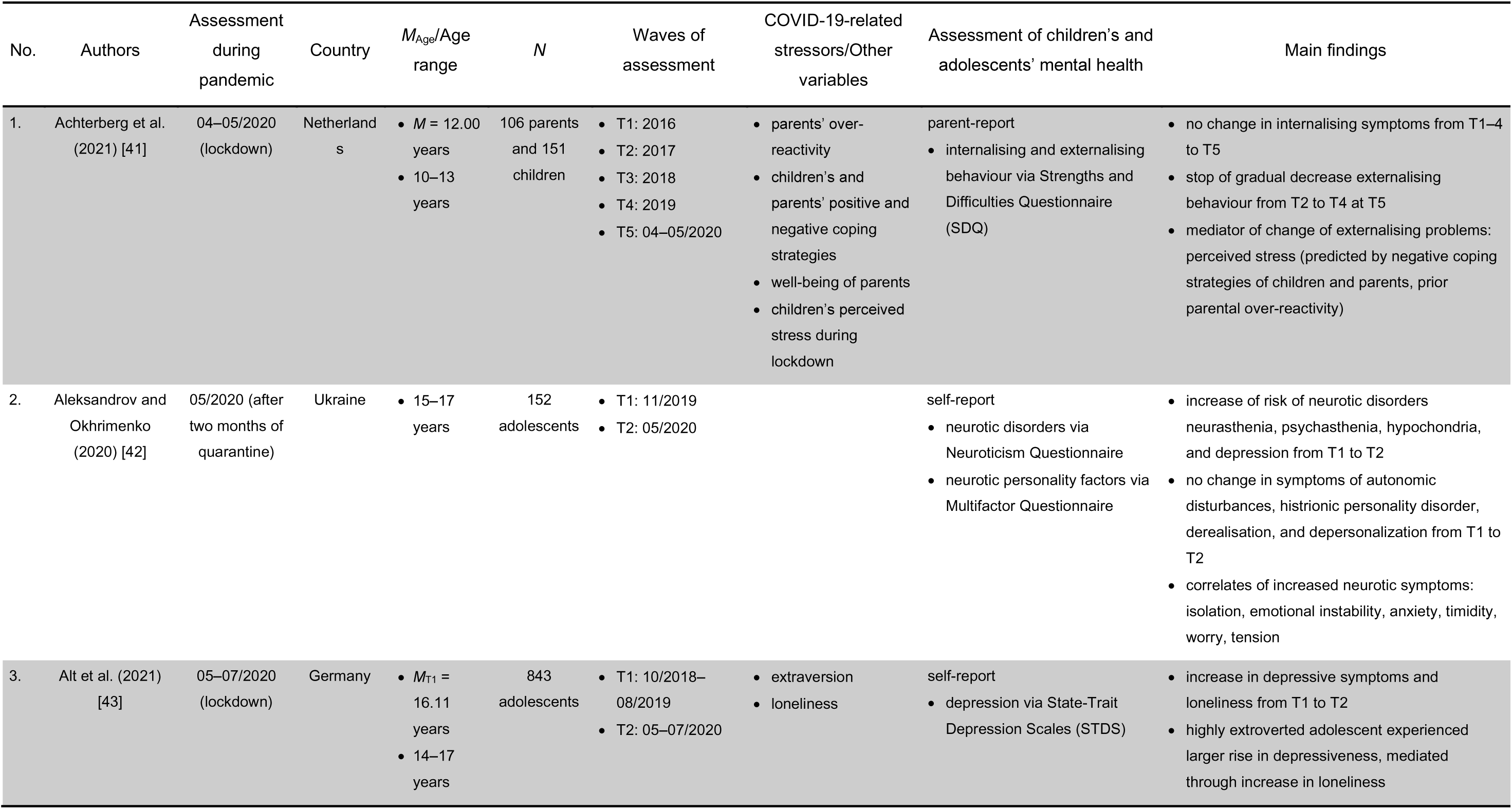
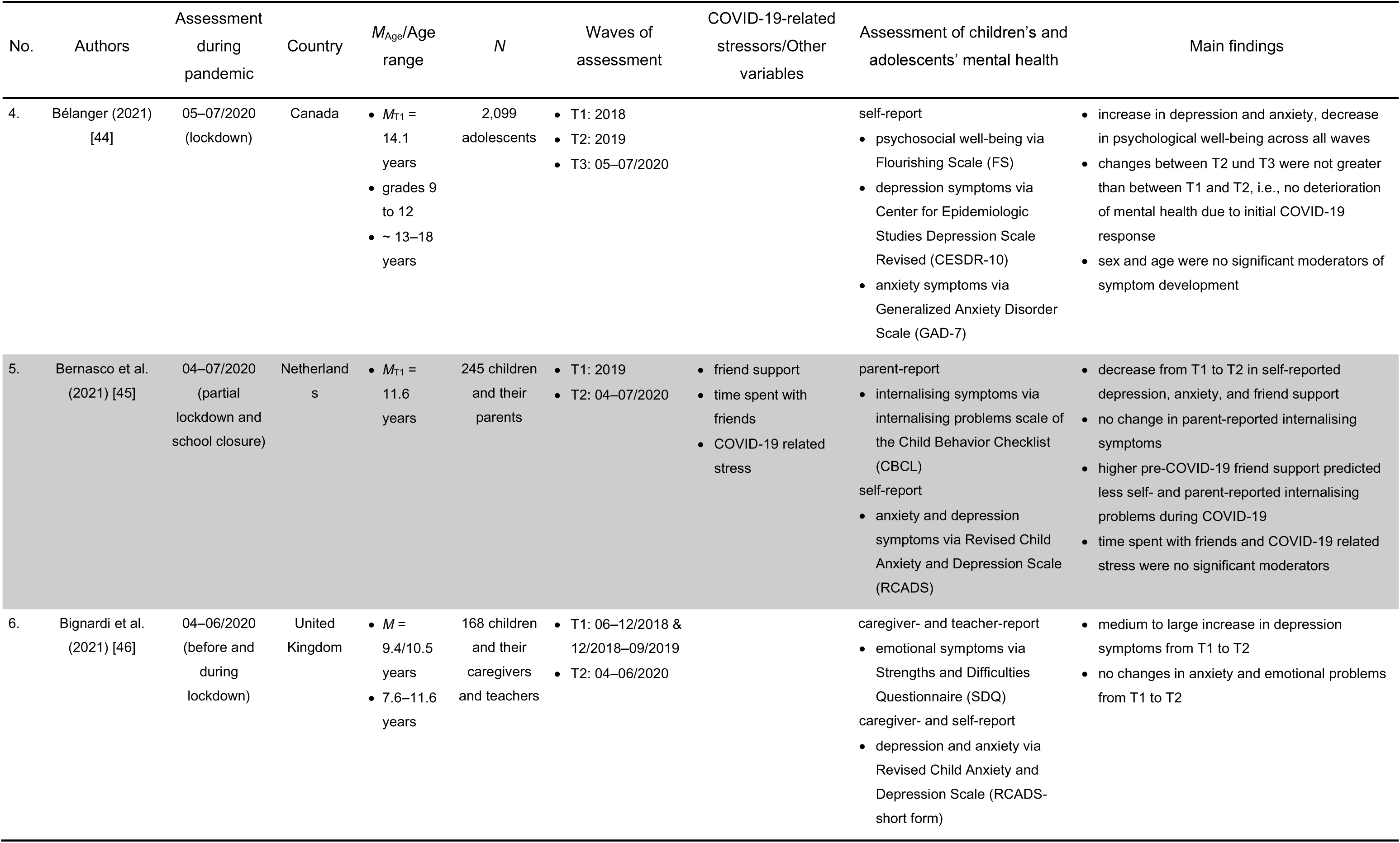
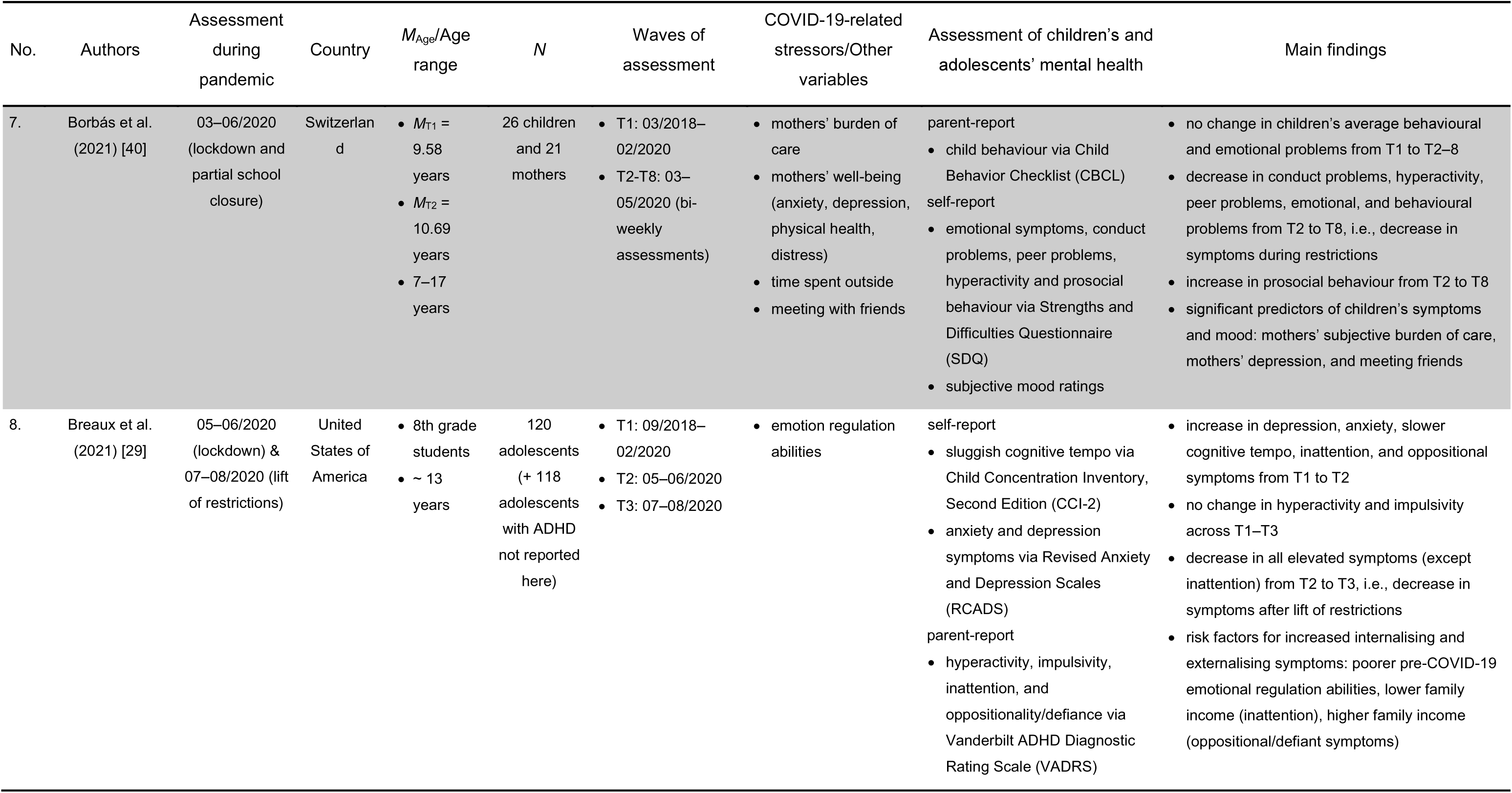
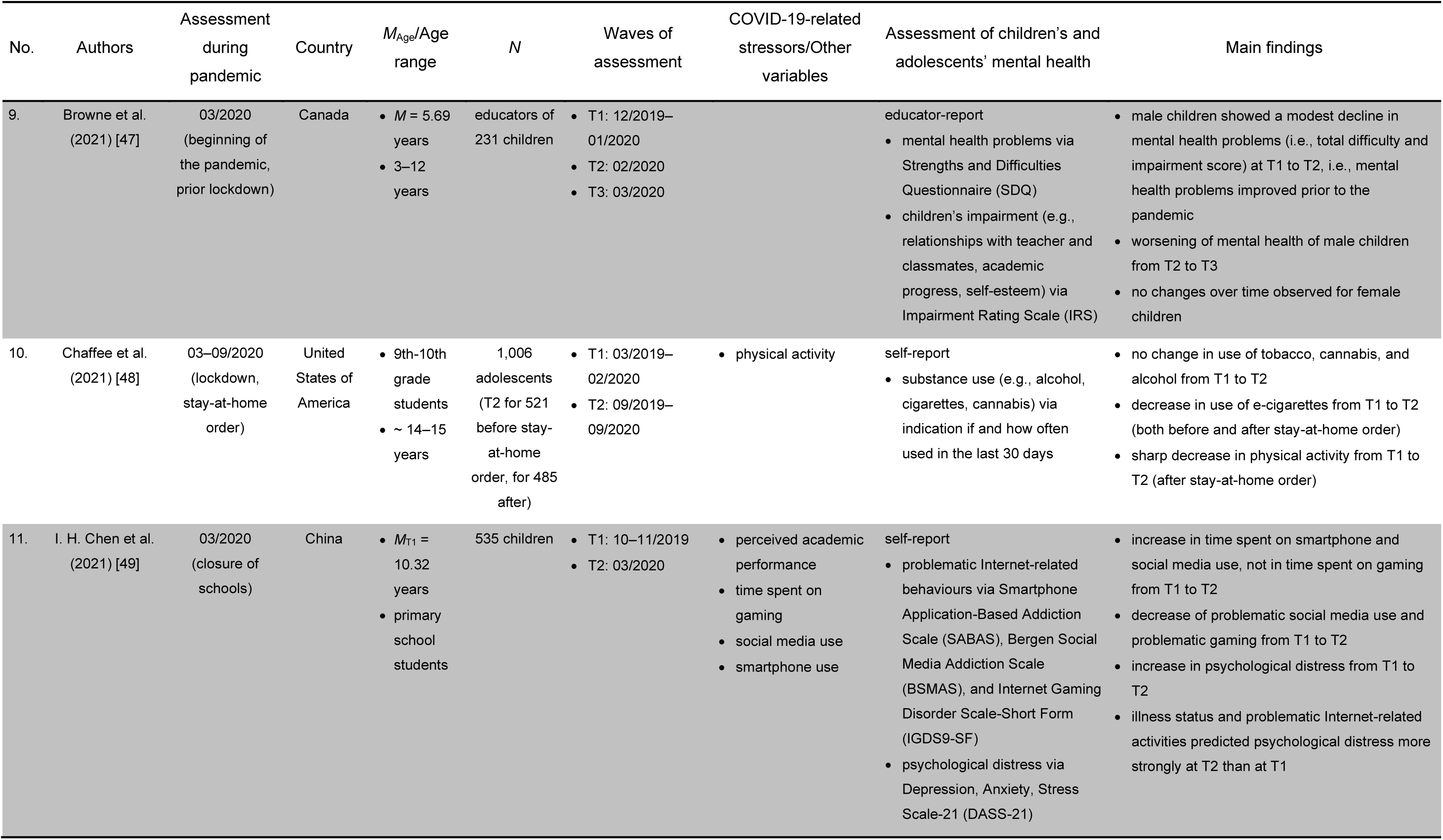
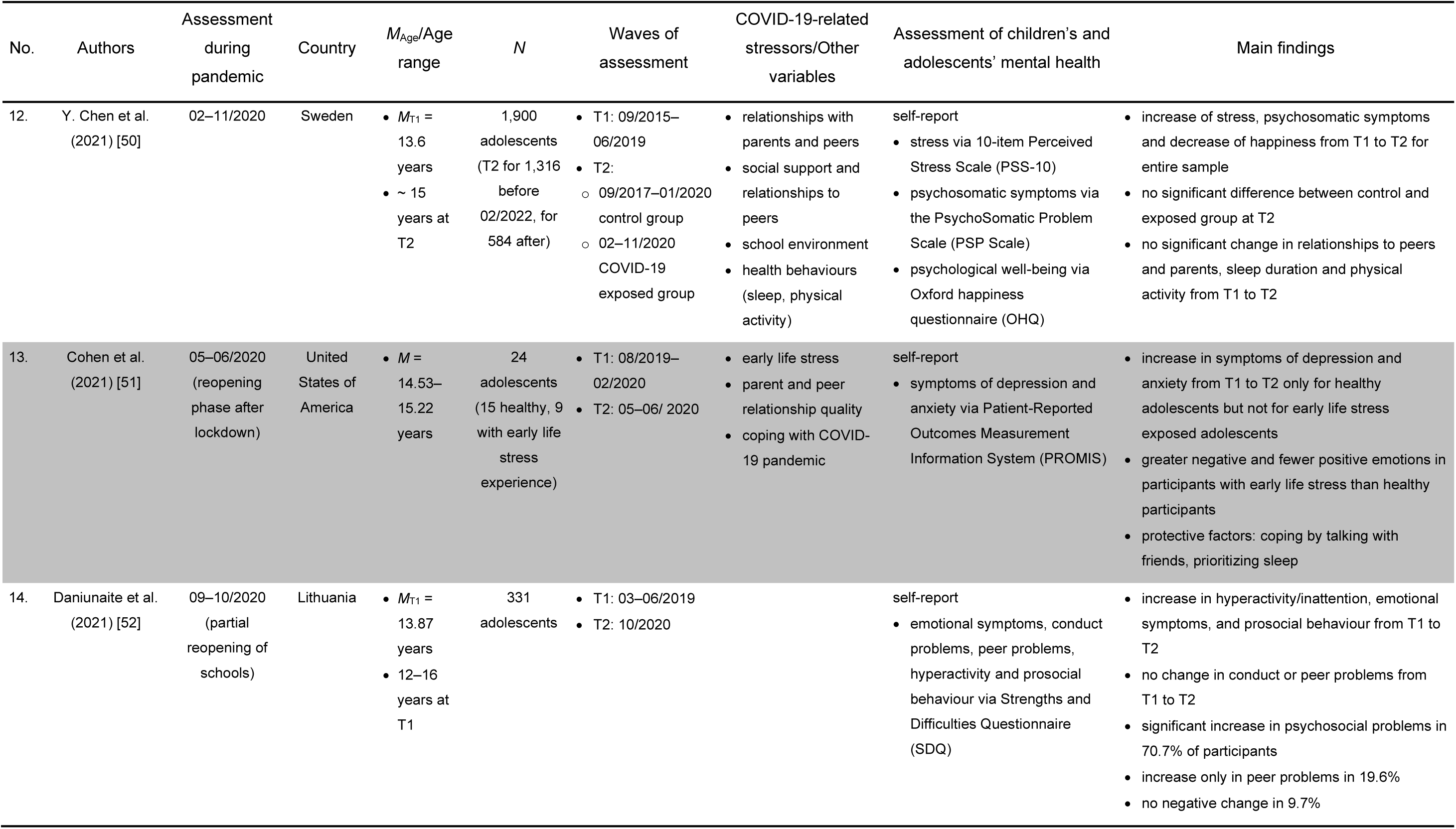
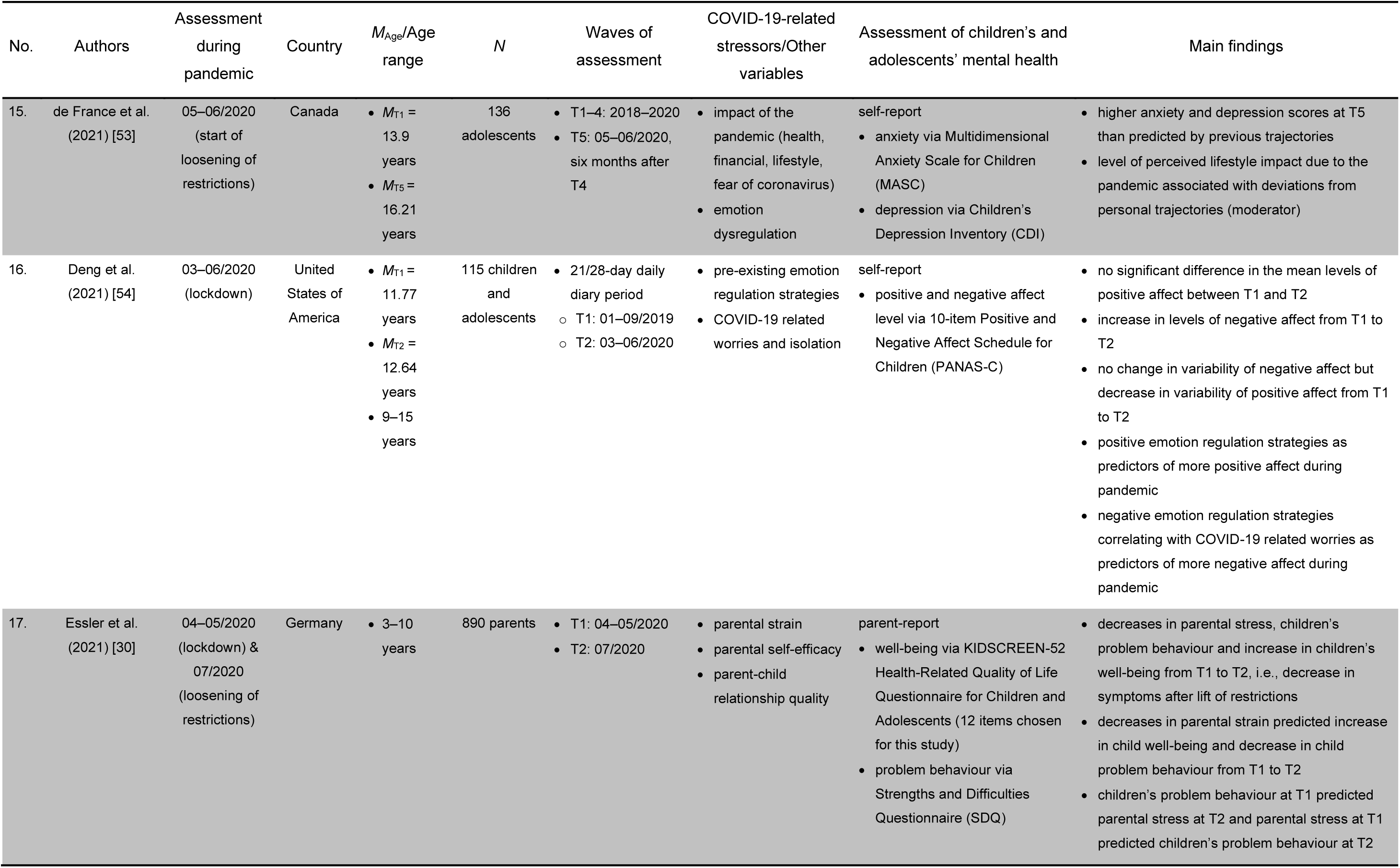
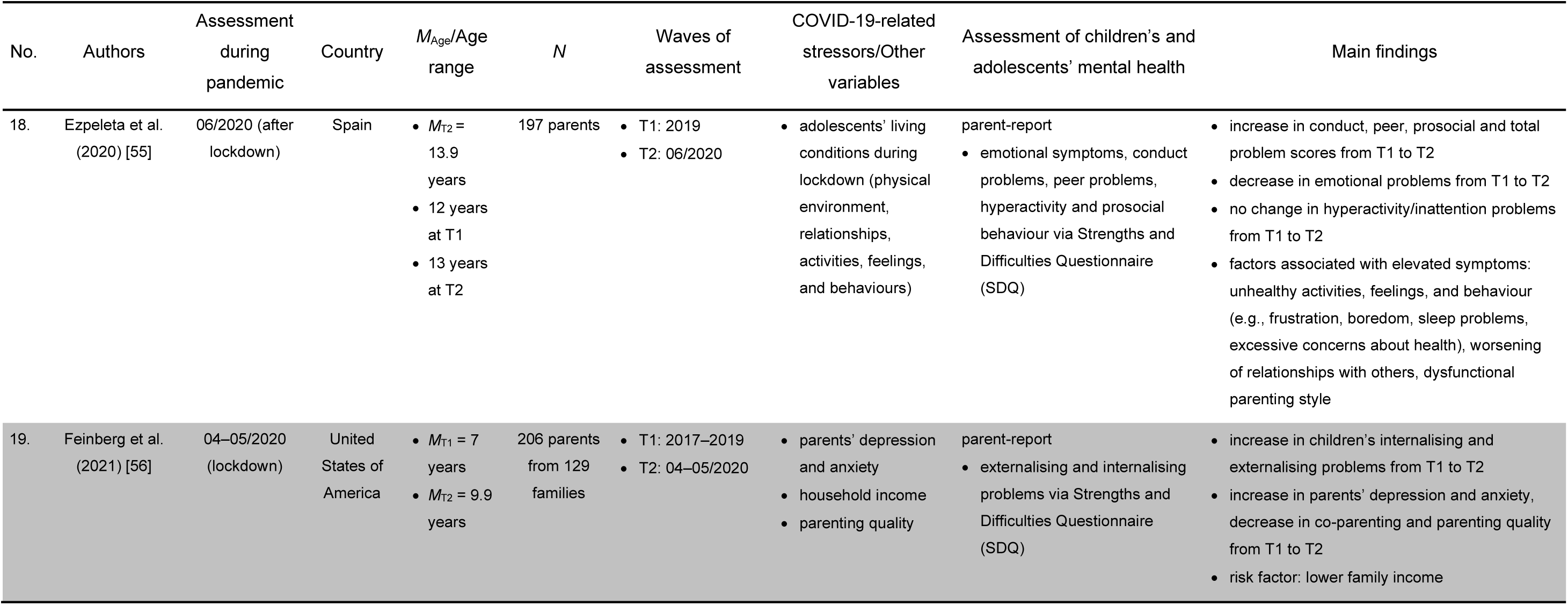
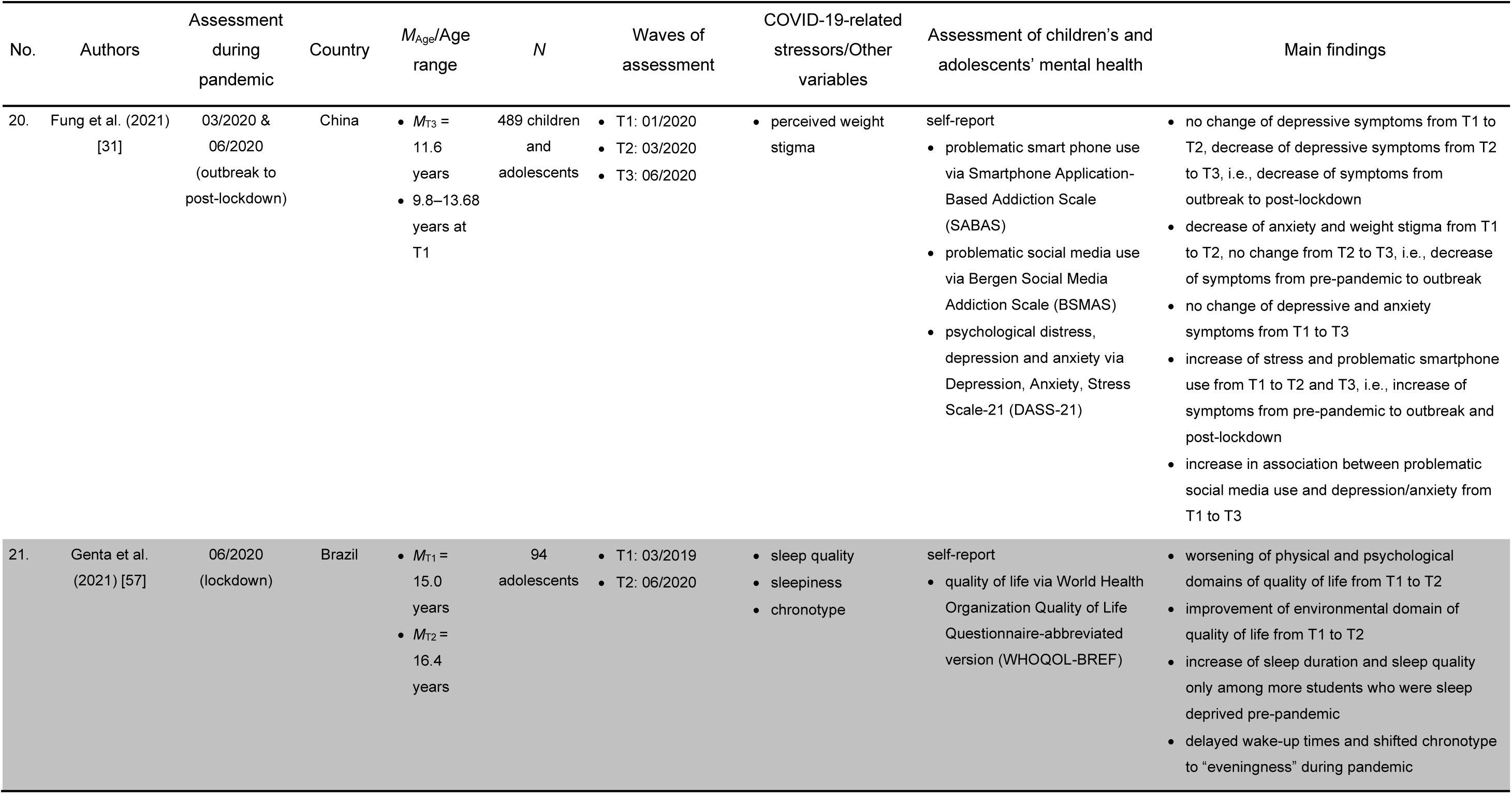
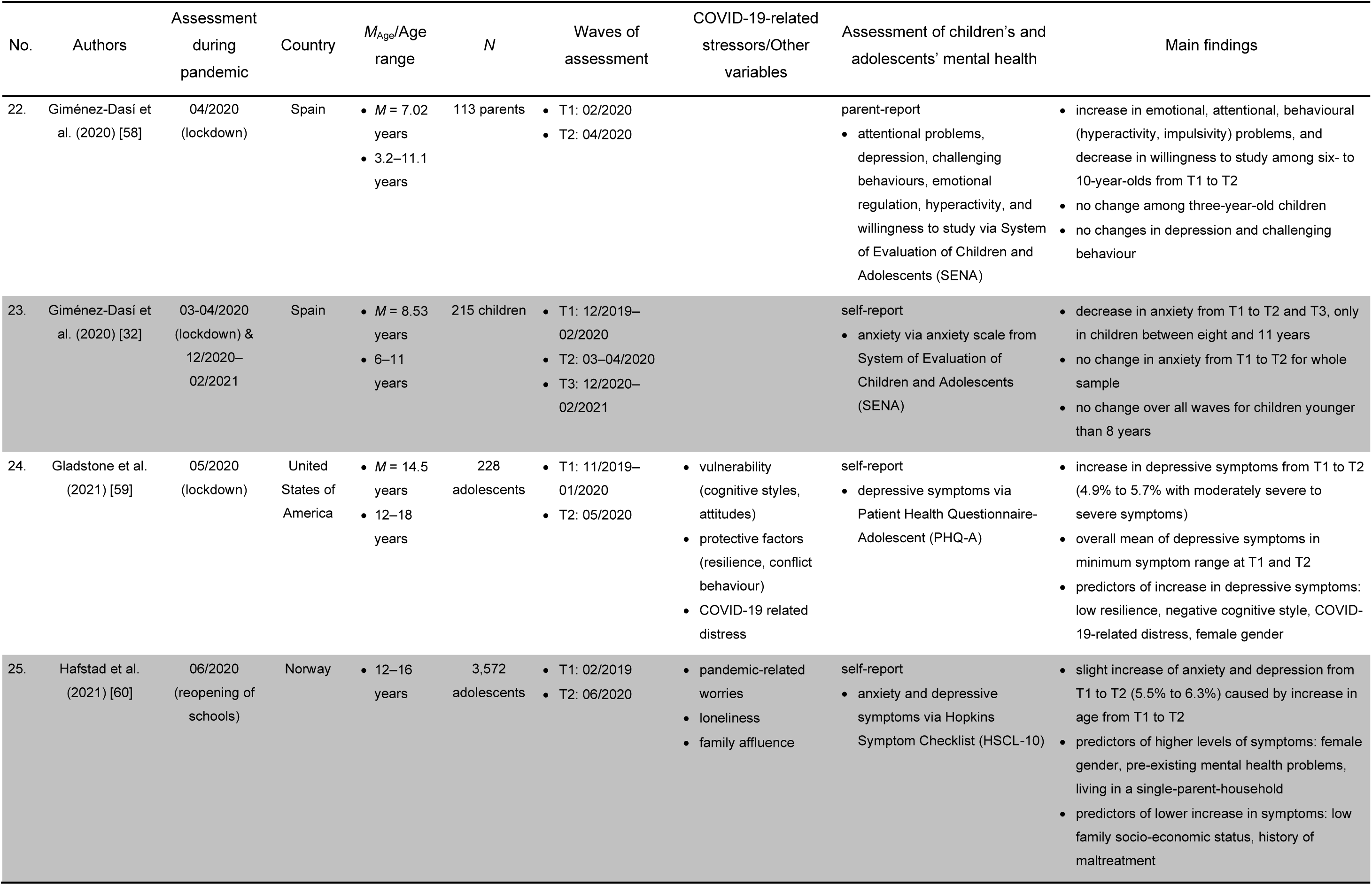
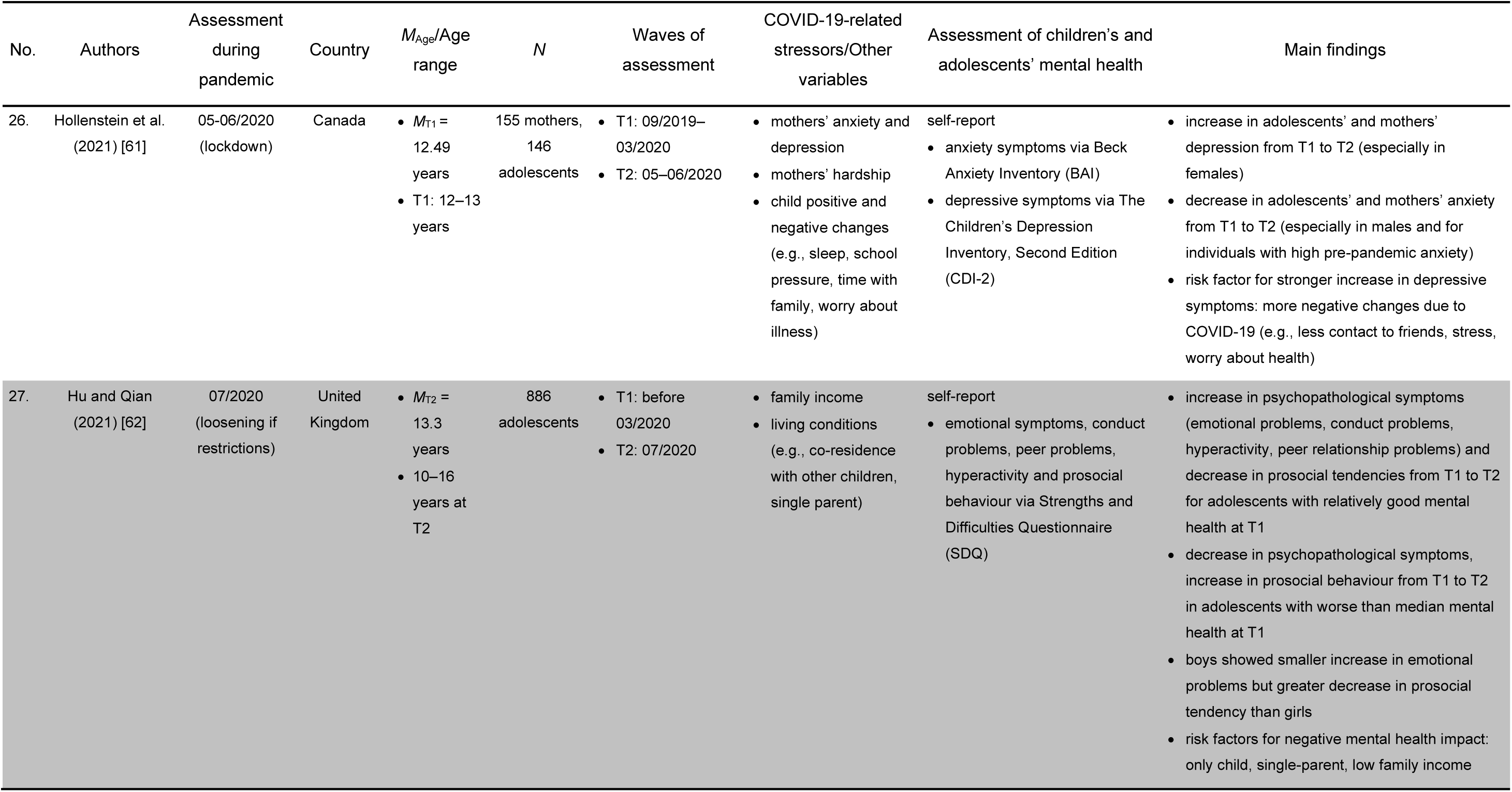
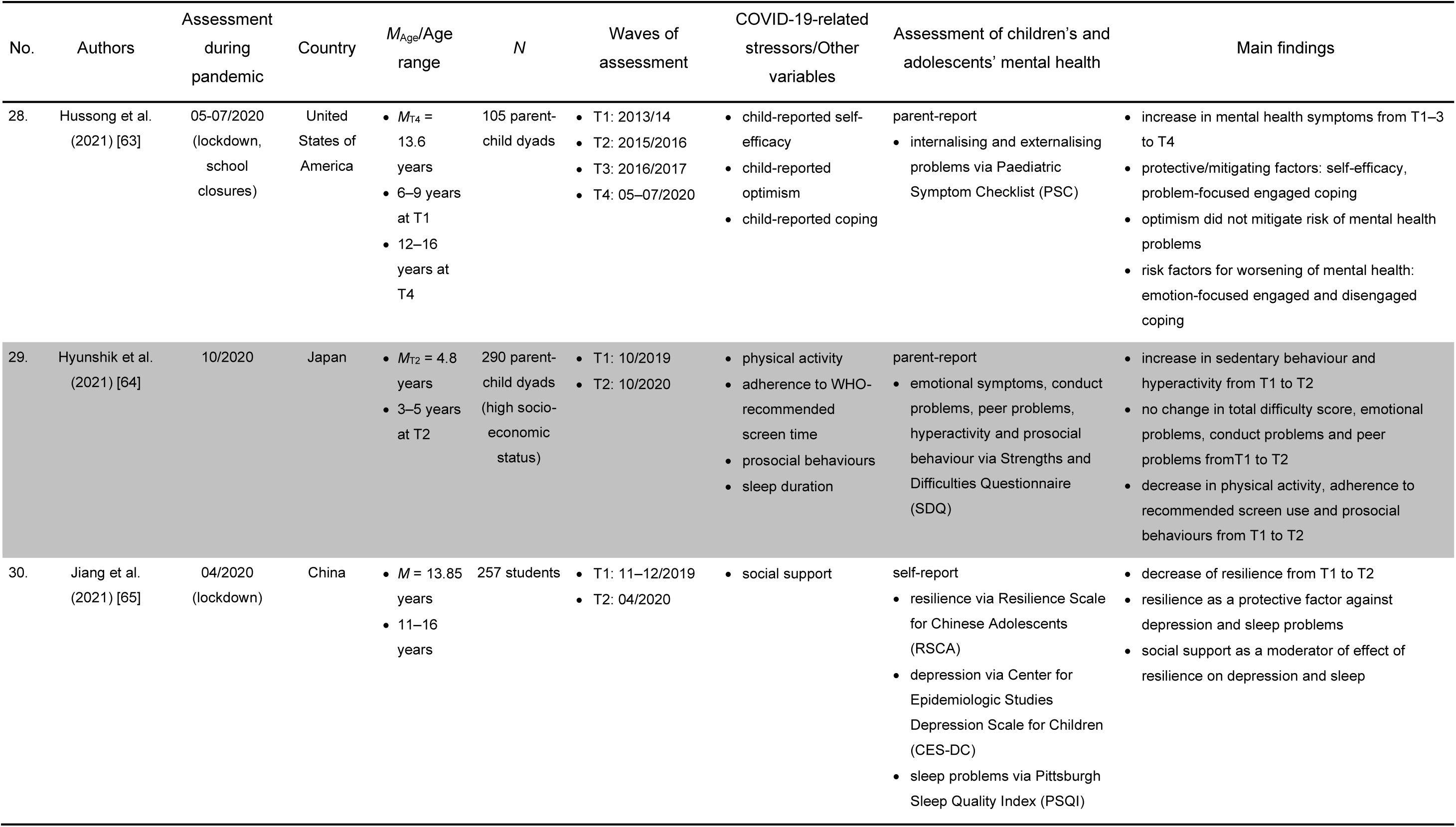
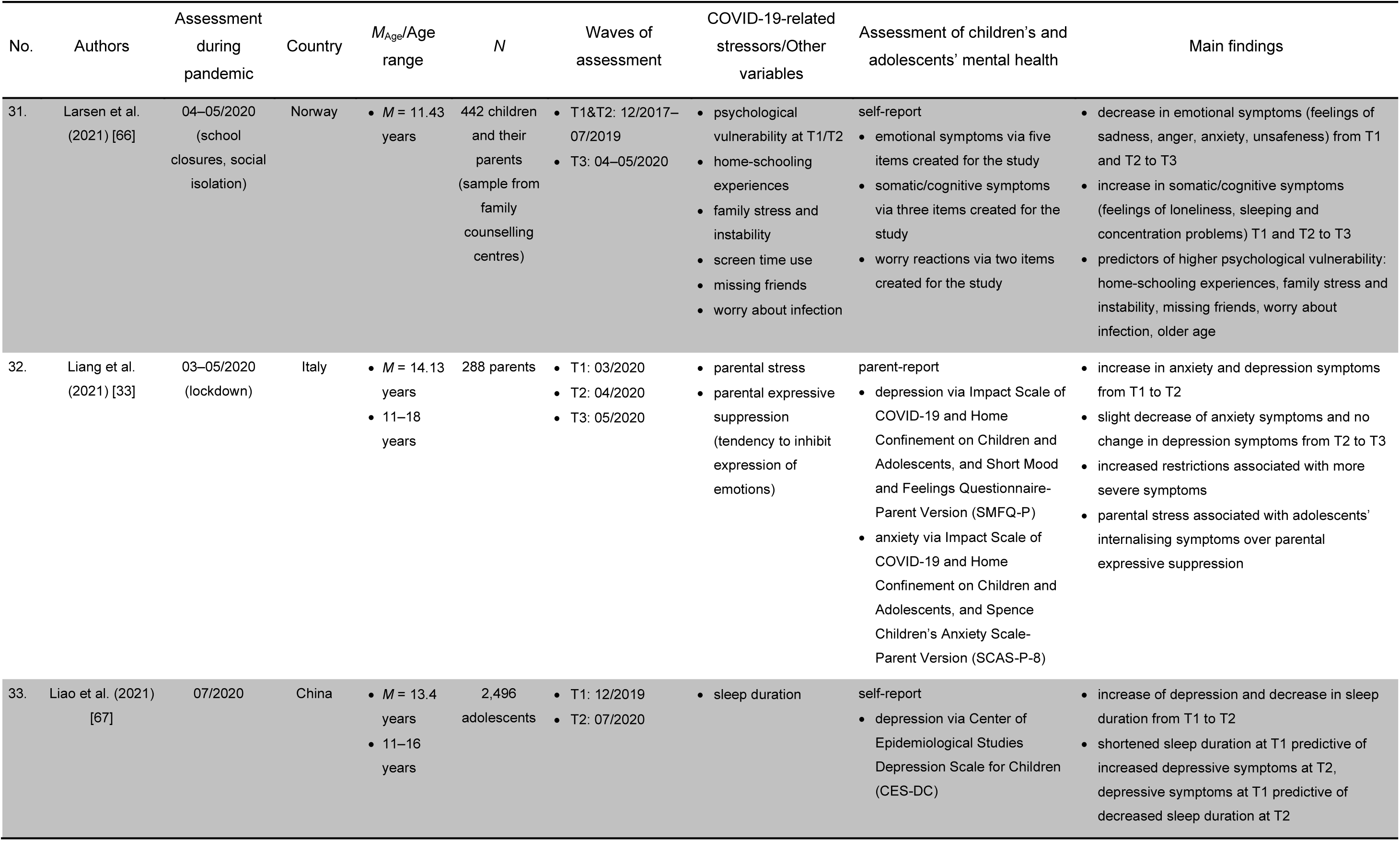
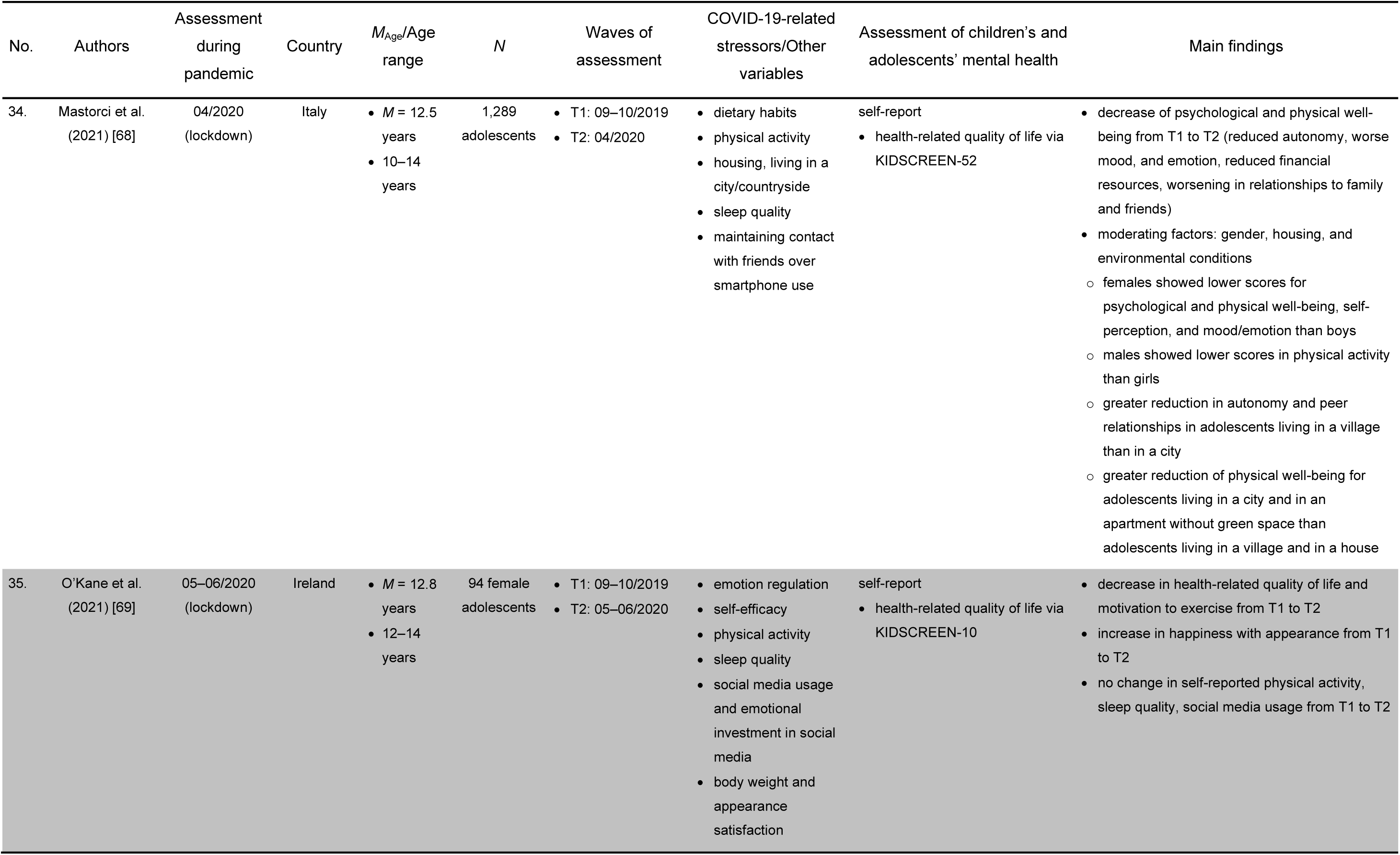
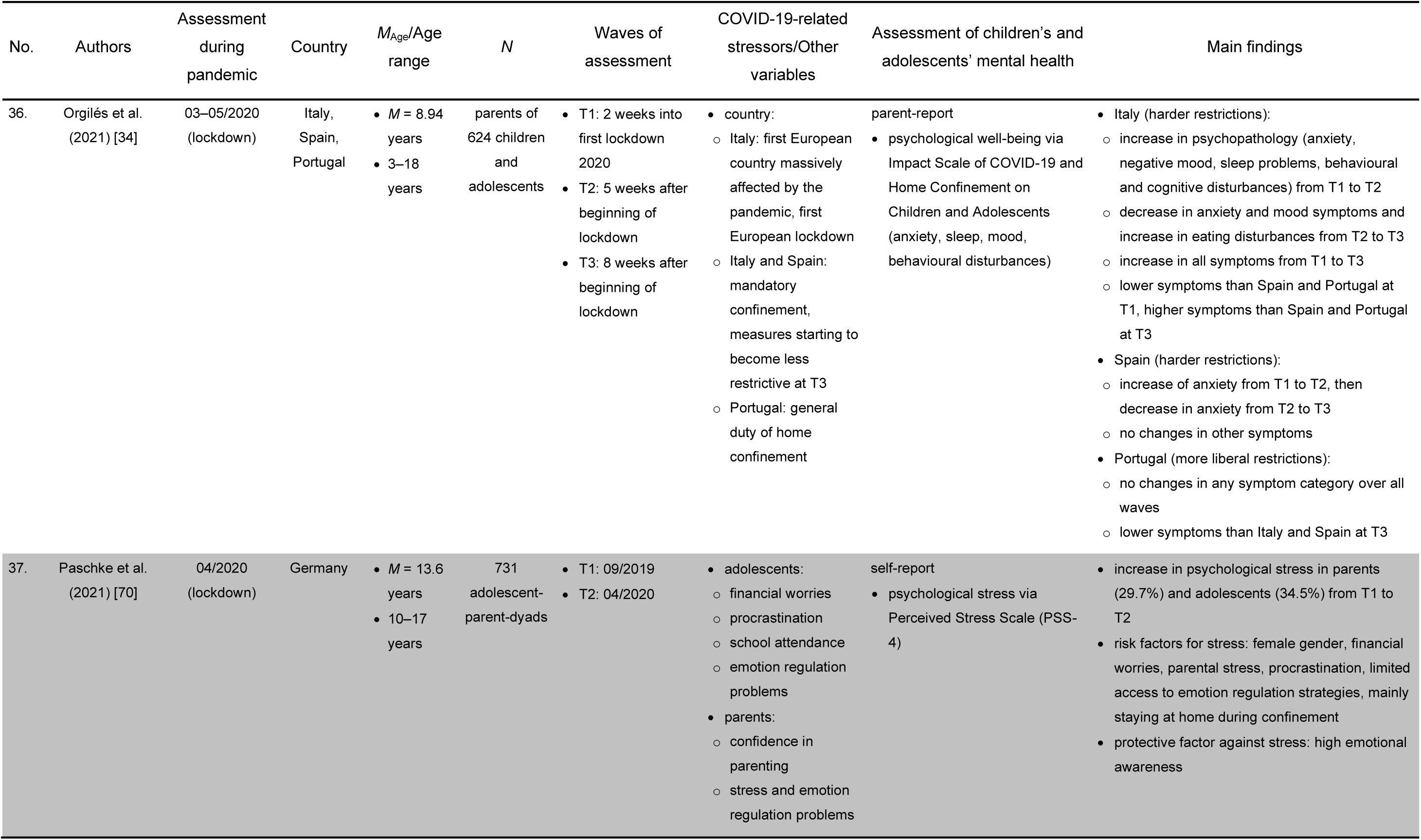
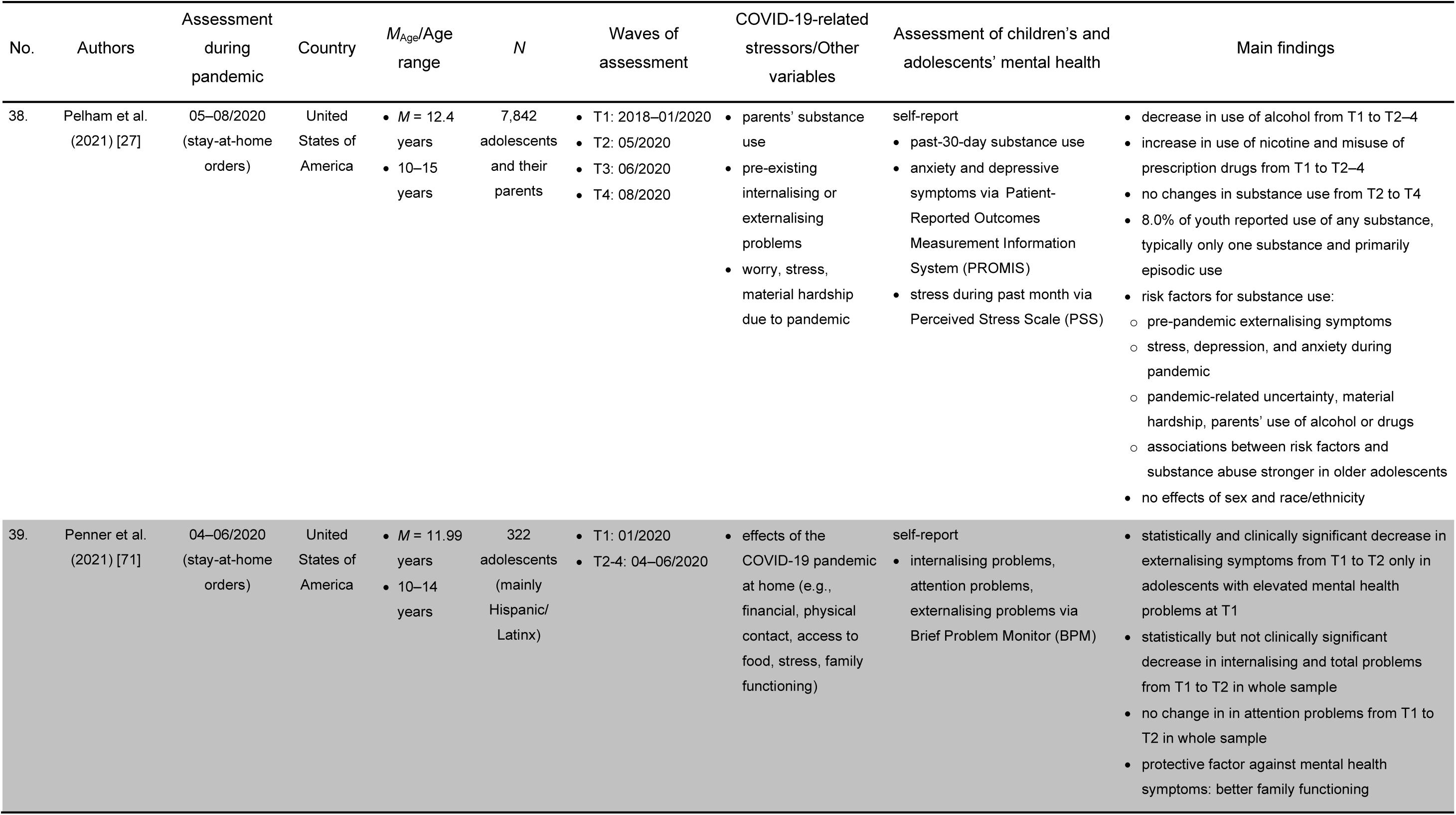
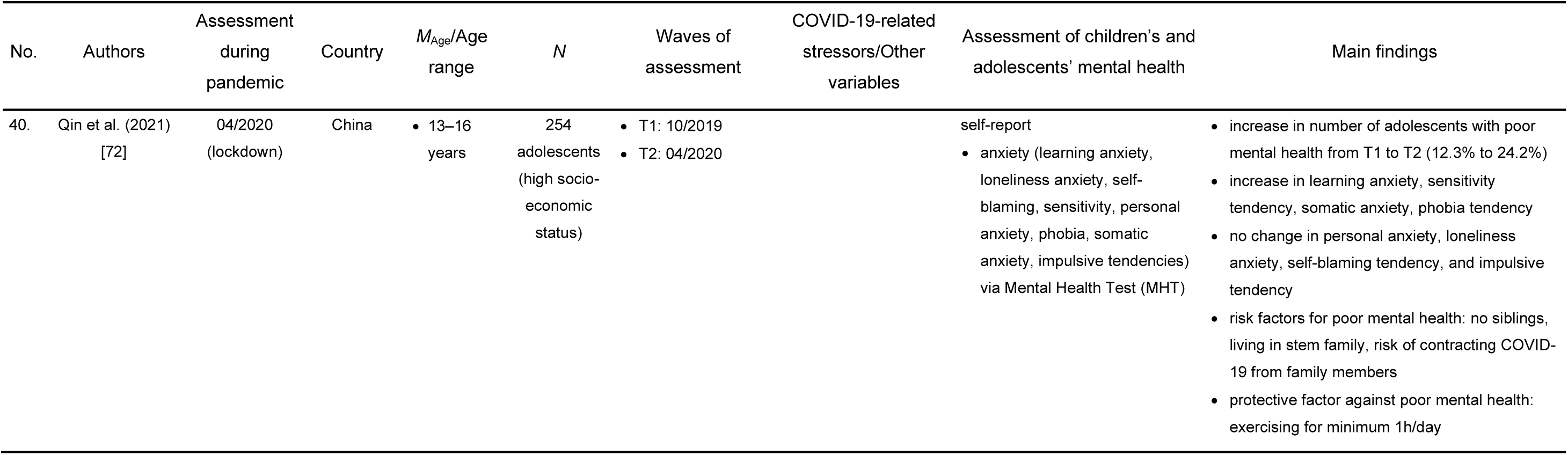
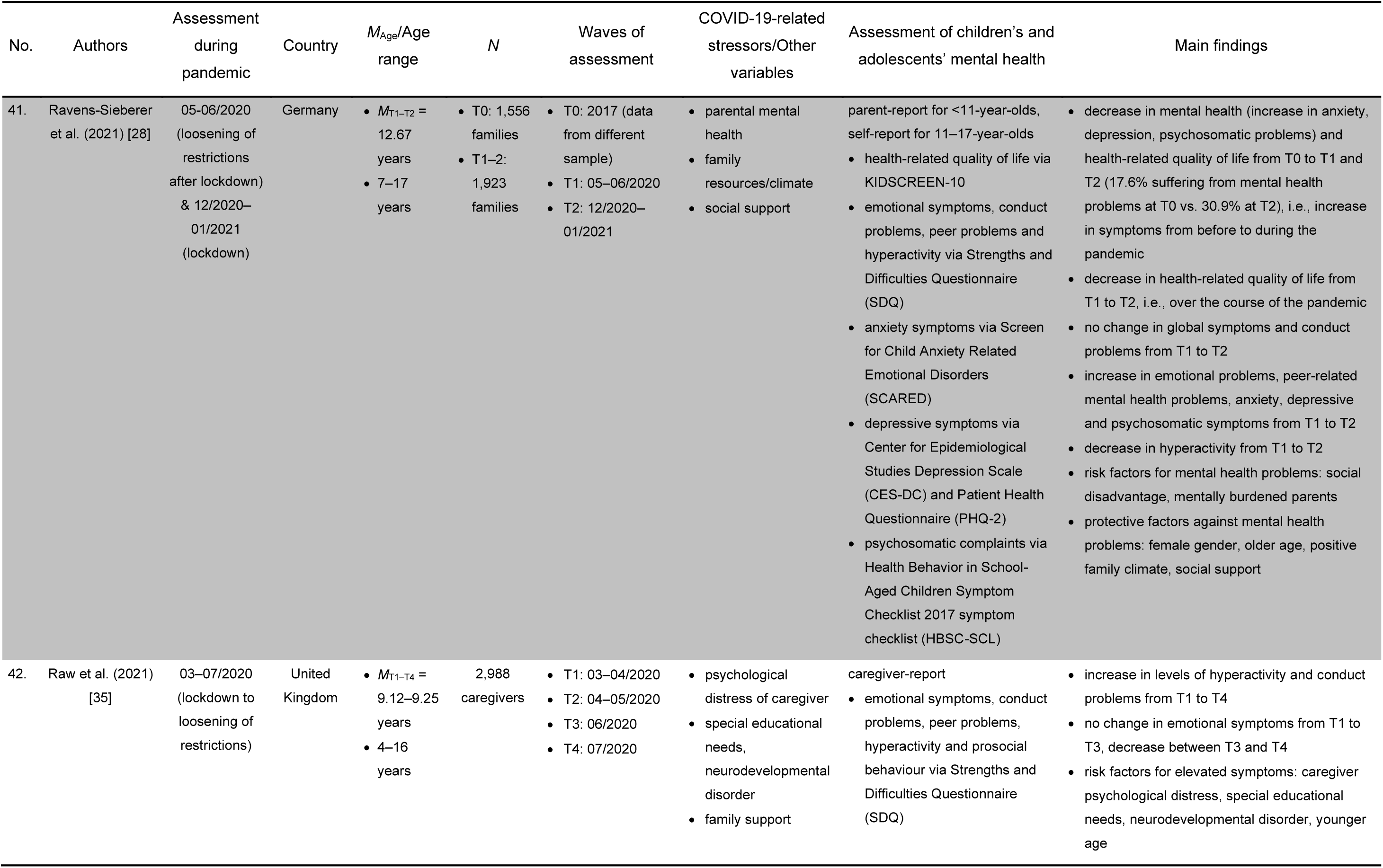
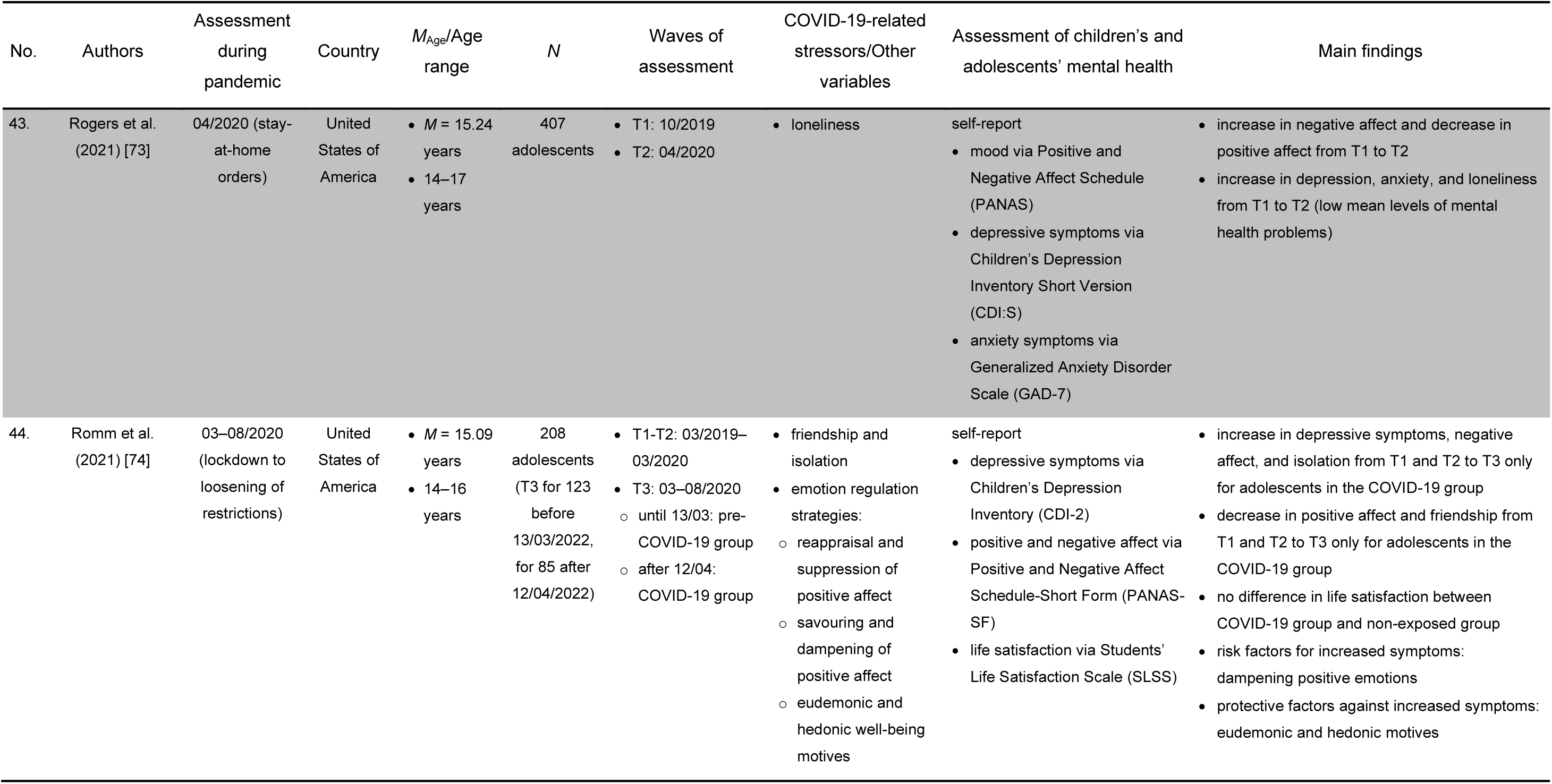
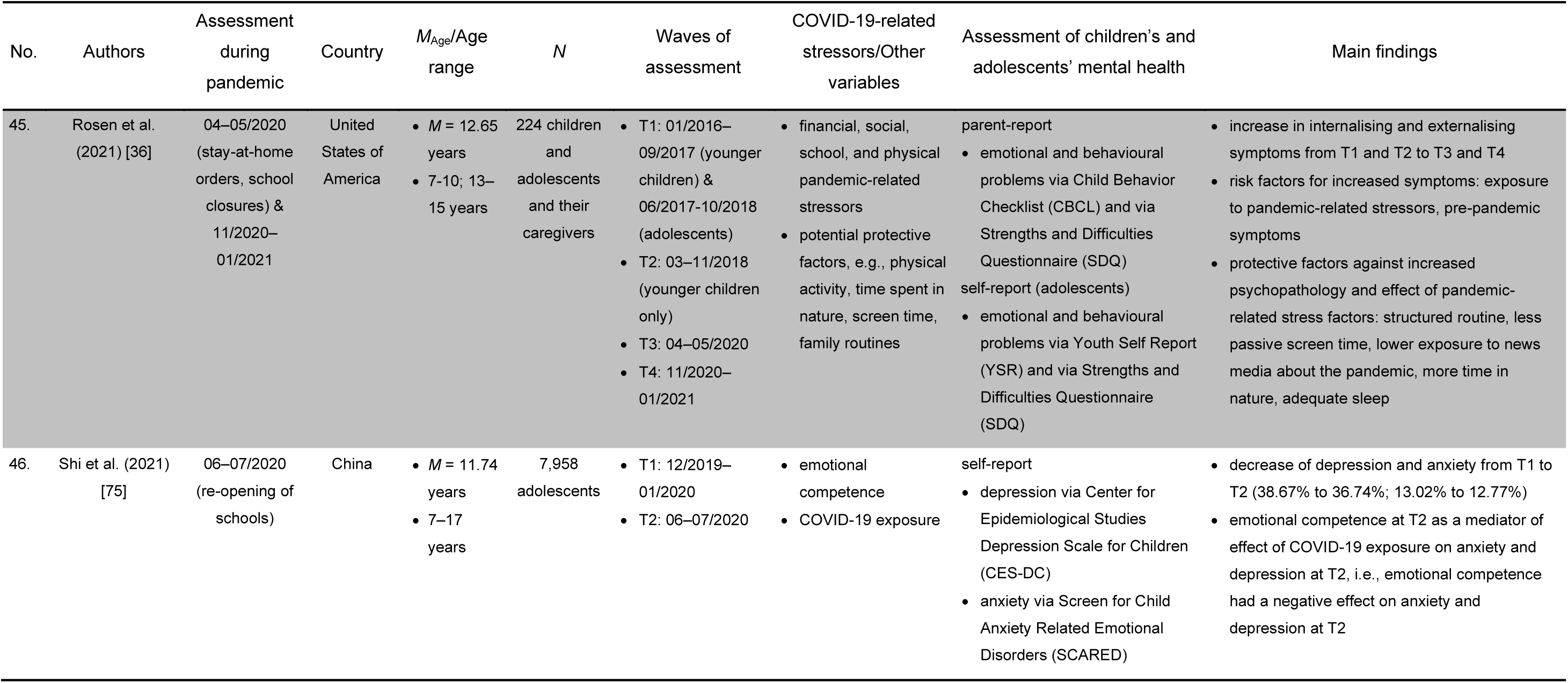
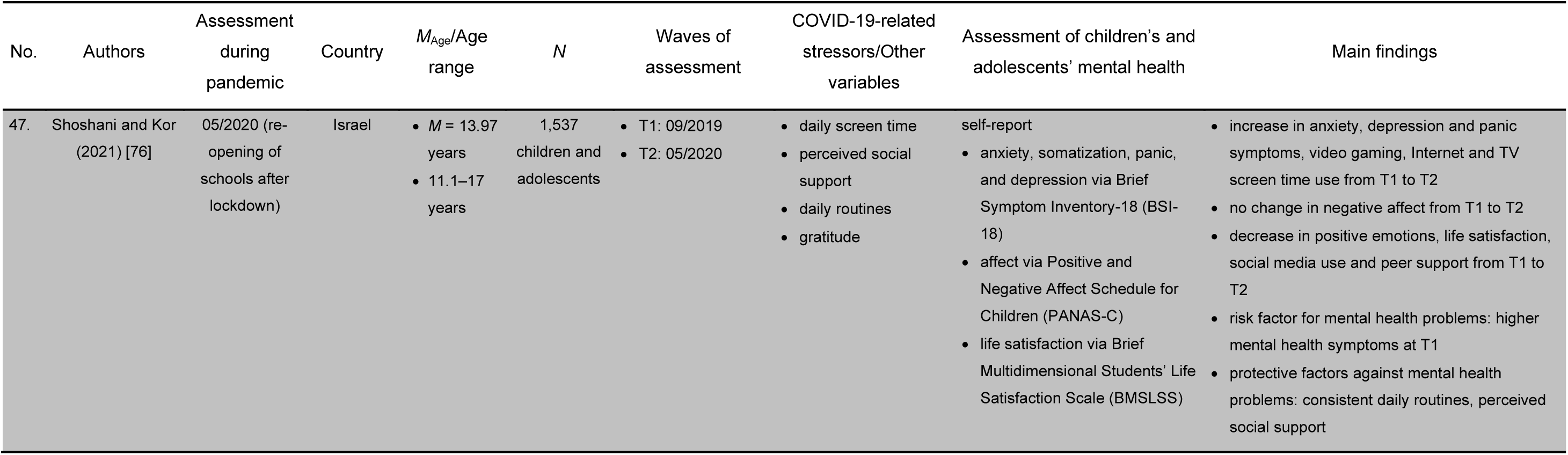
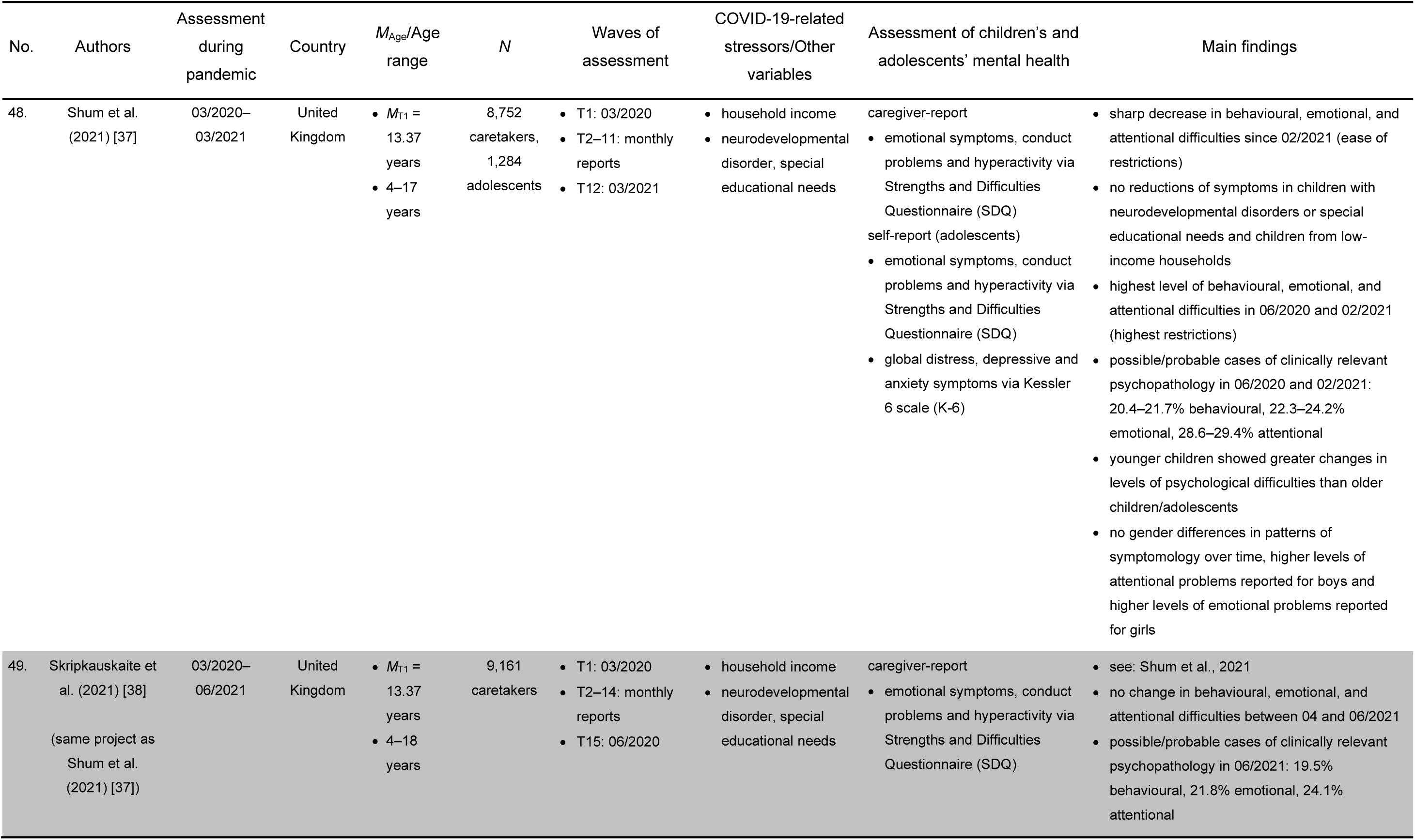
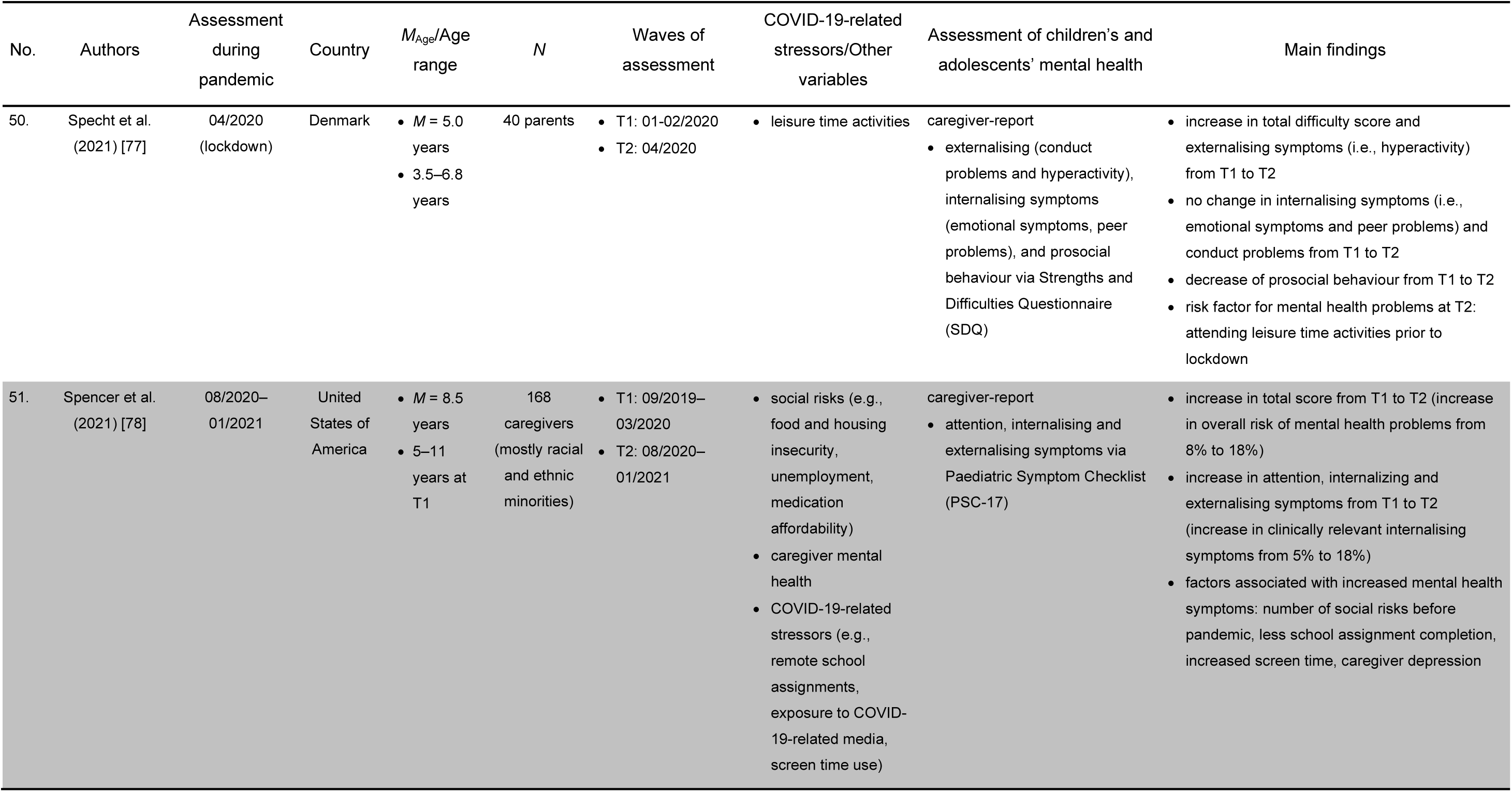
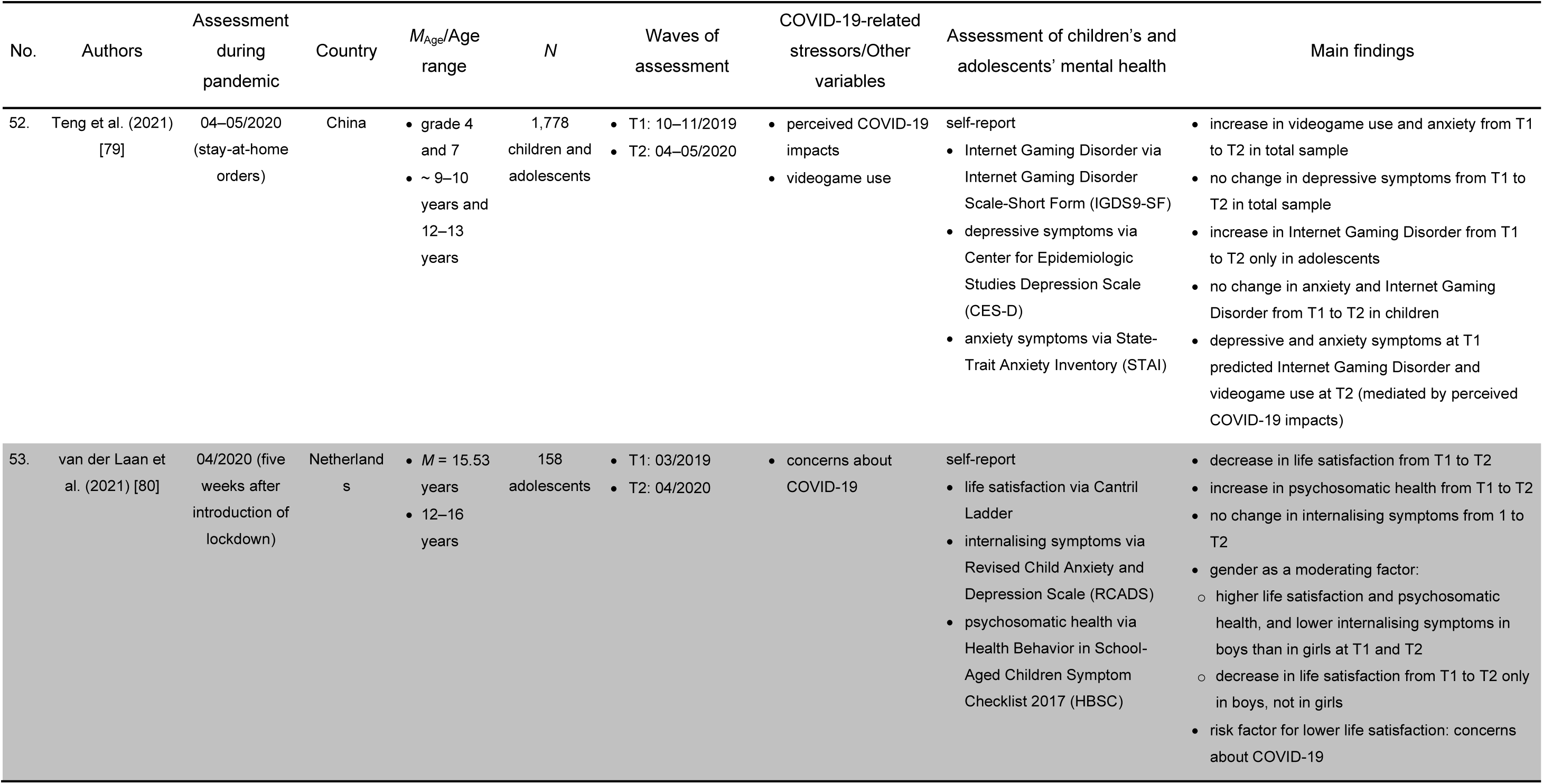
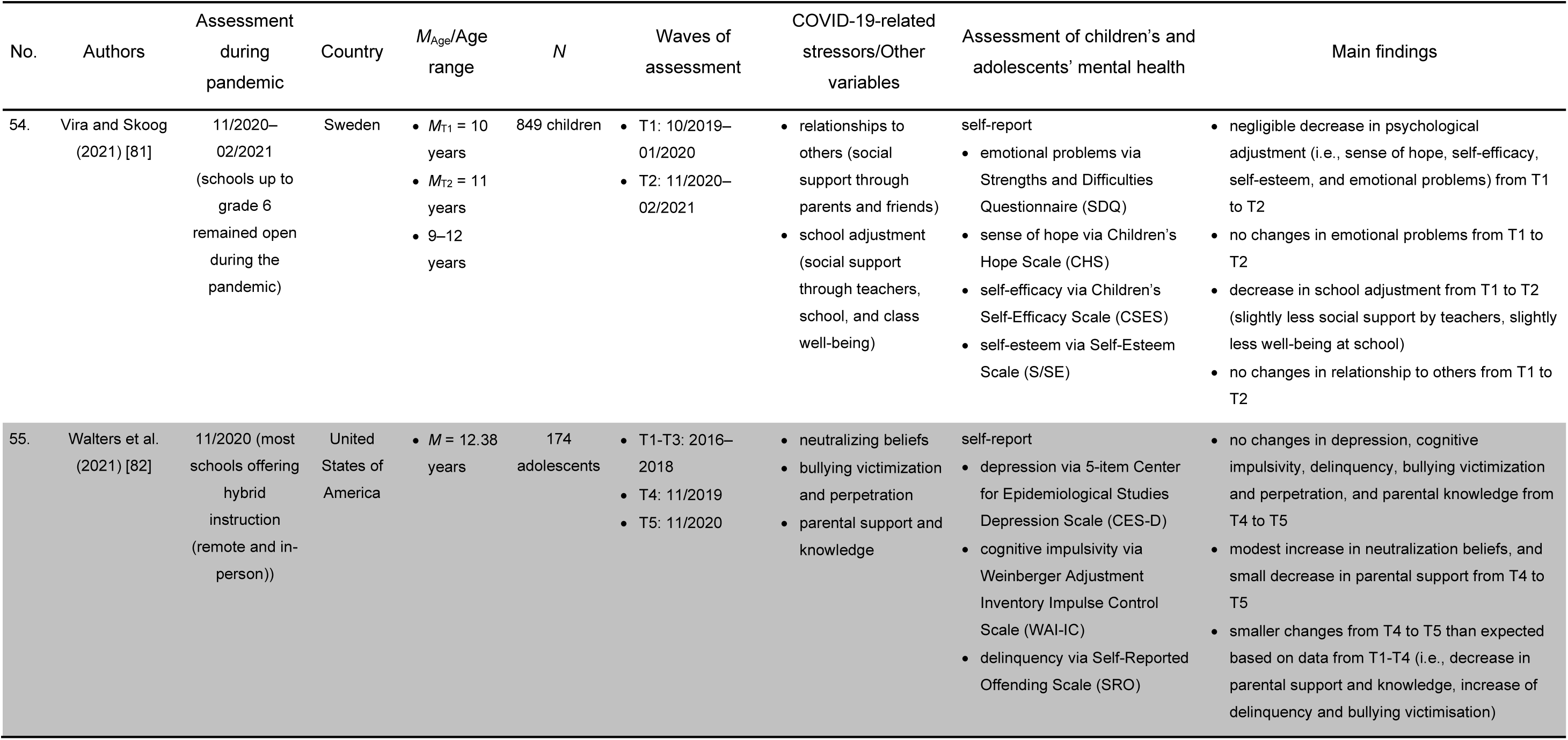
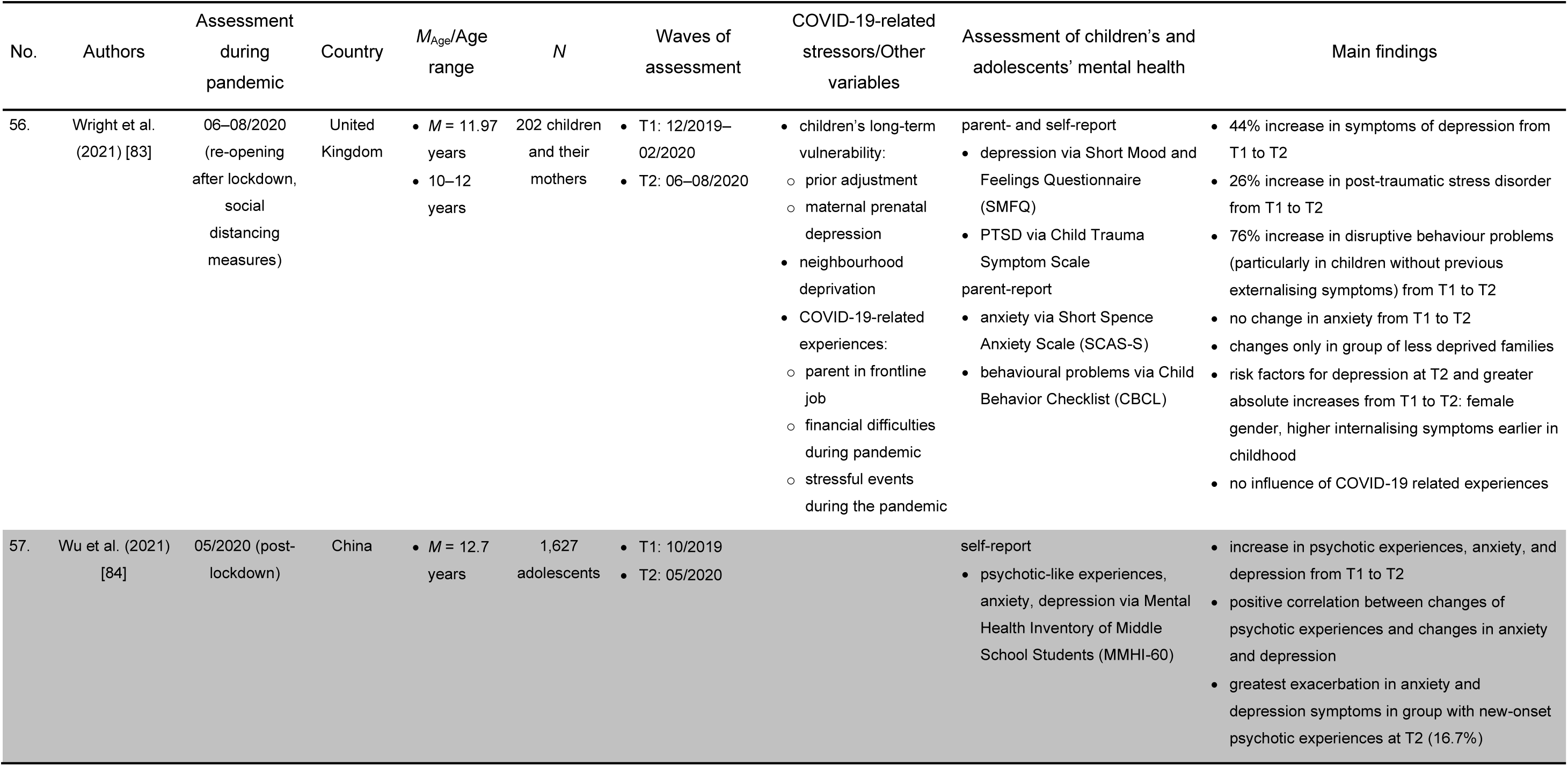
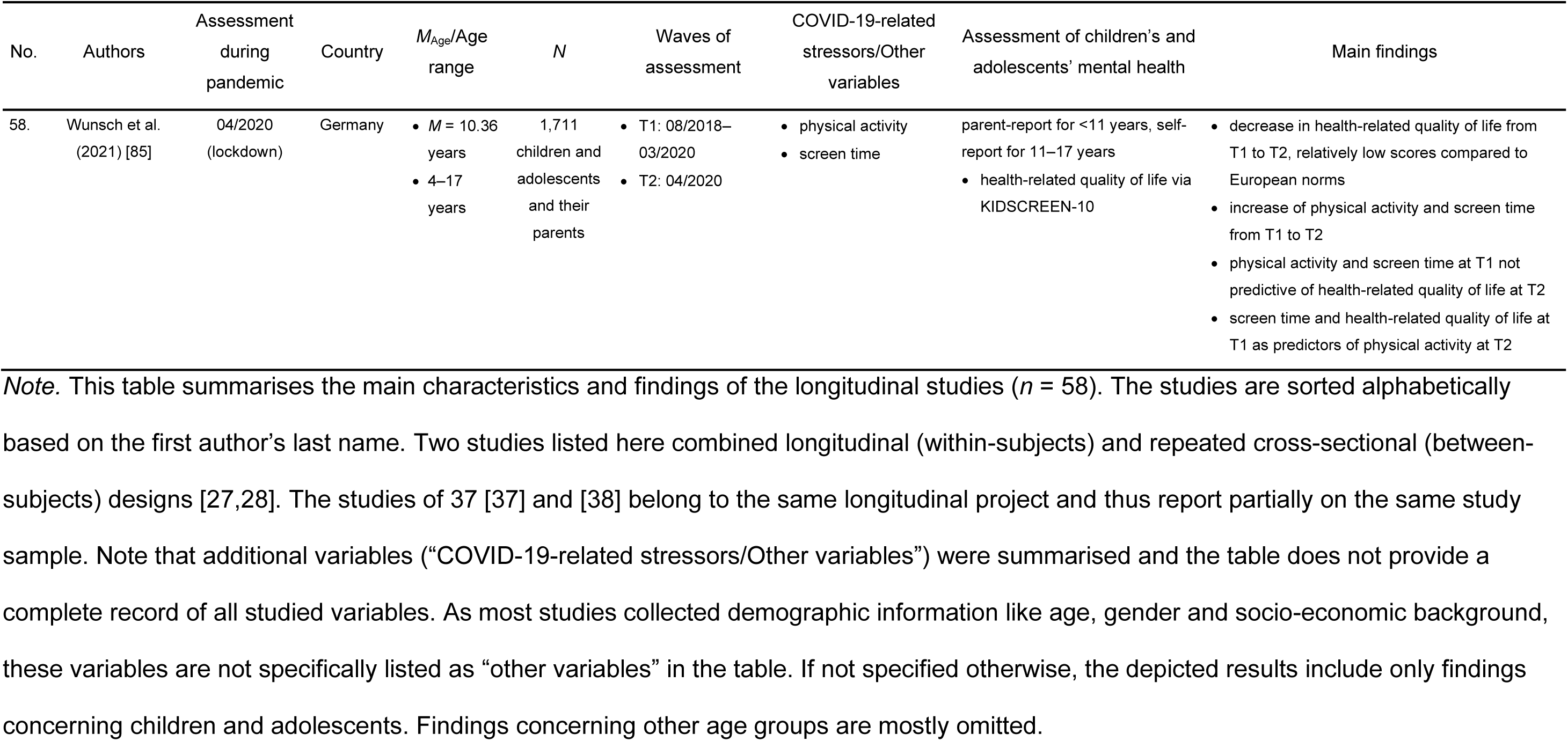
Overview of Longitudinal Studies (Same Sample).

**Table 2.**
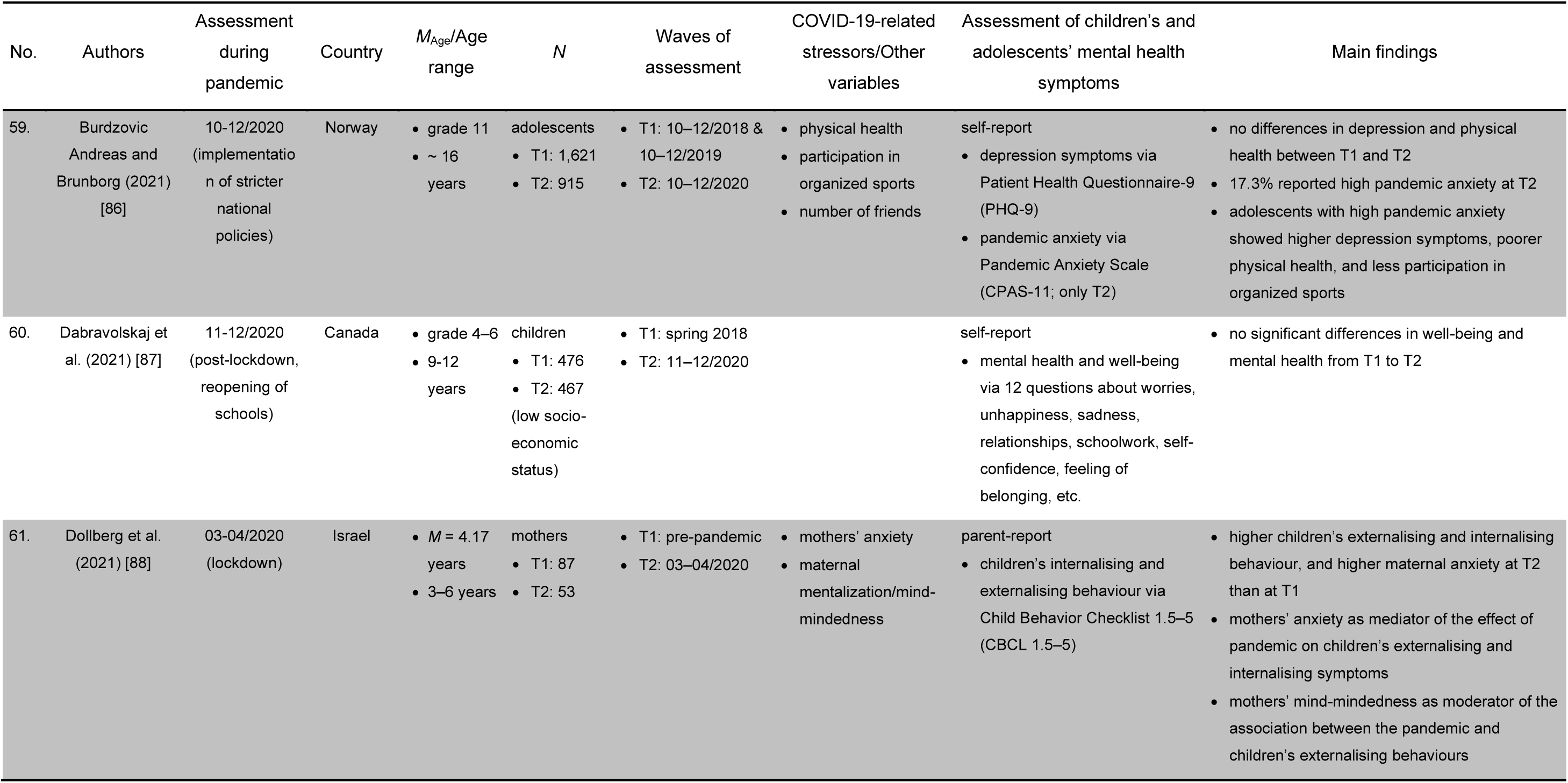
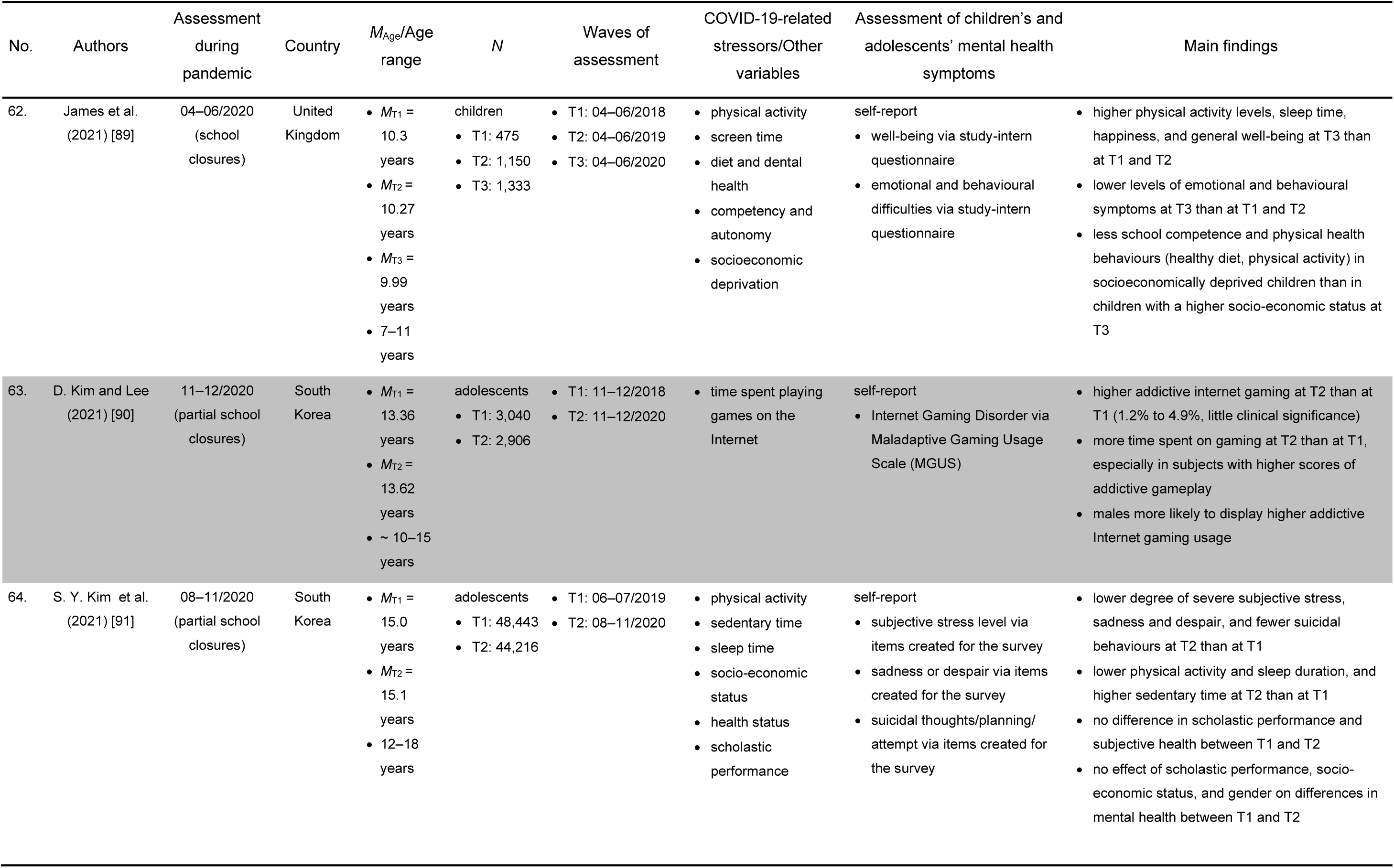
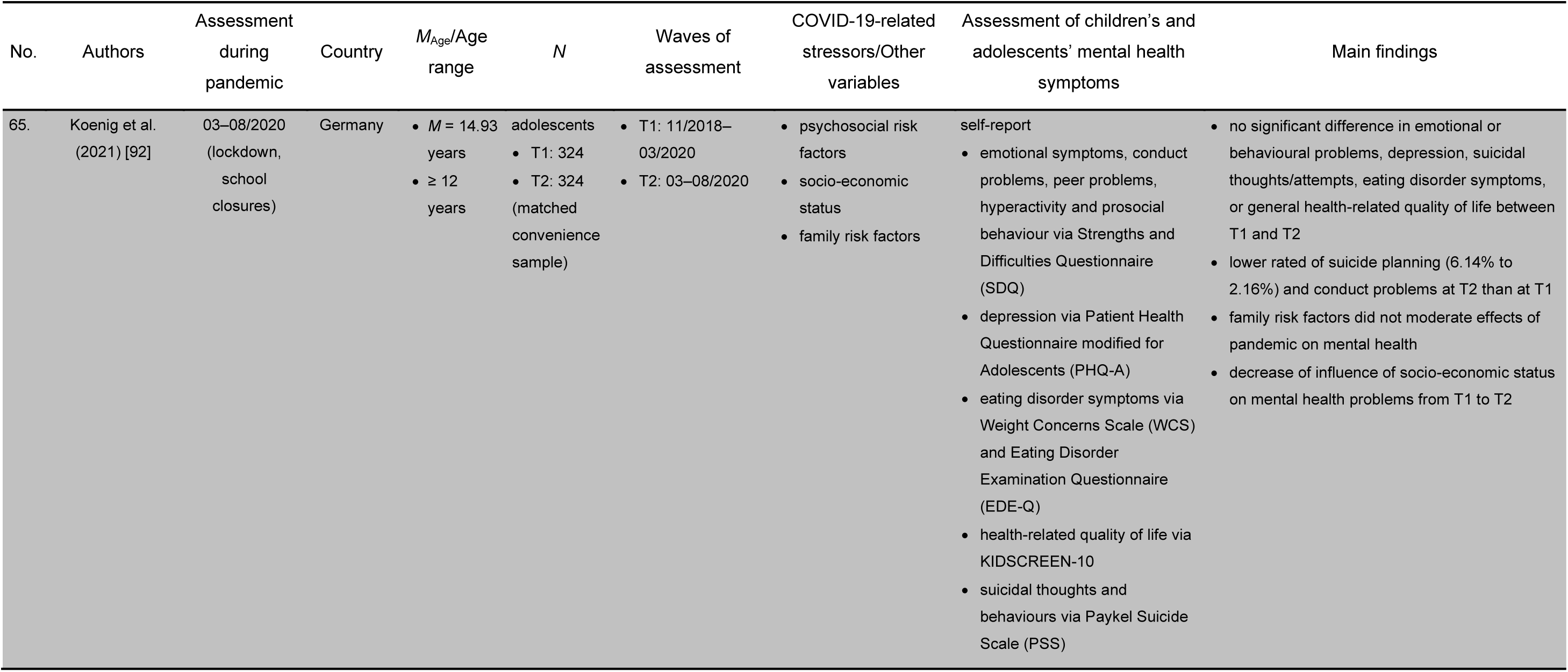
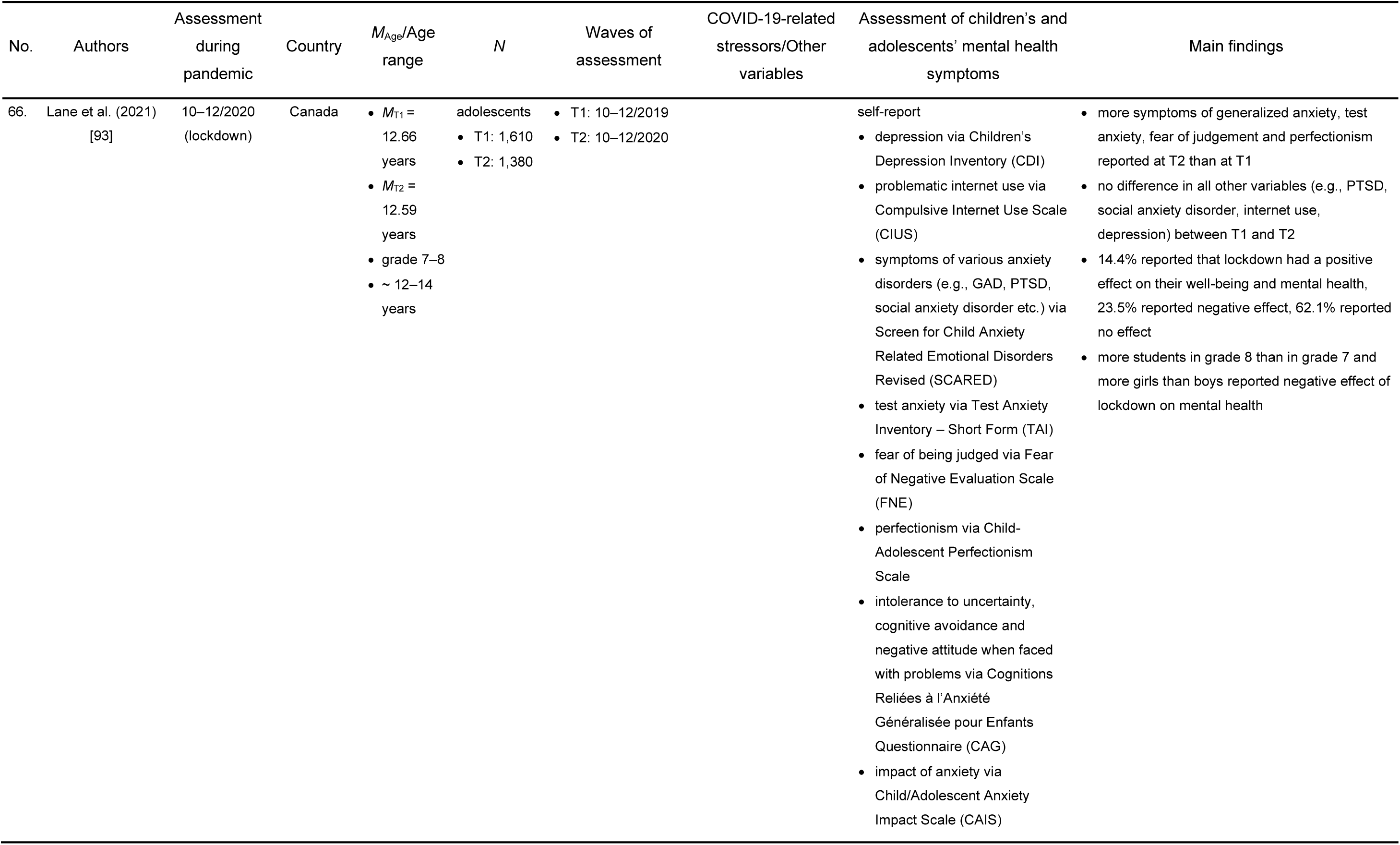
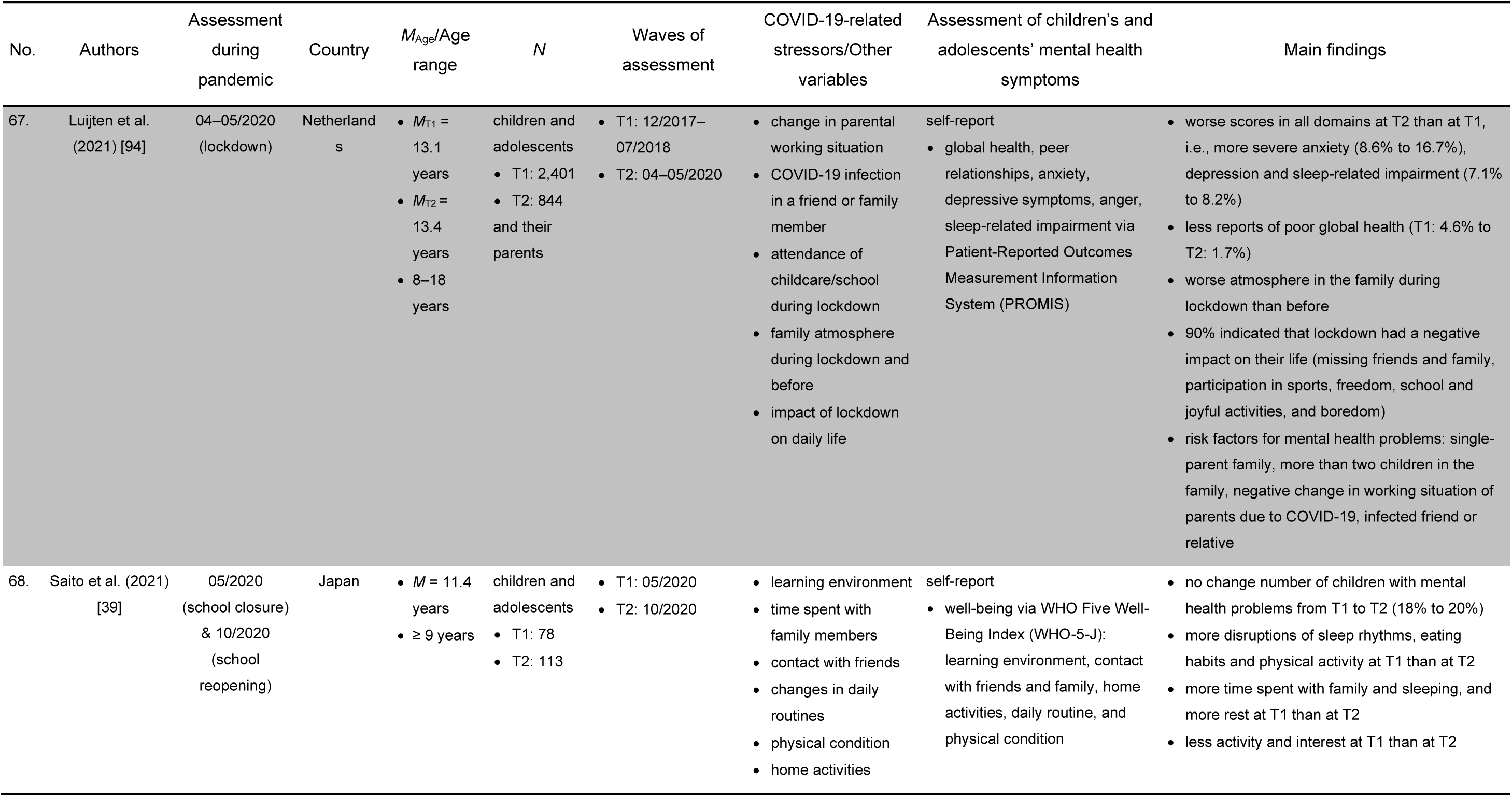
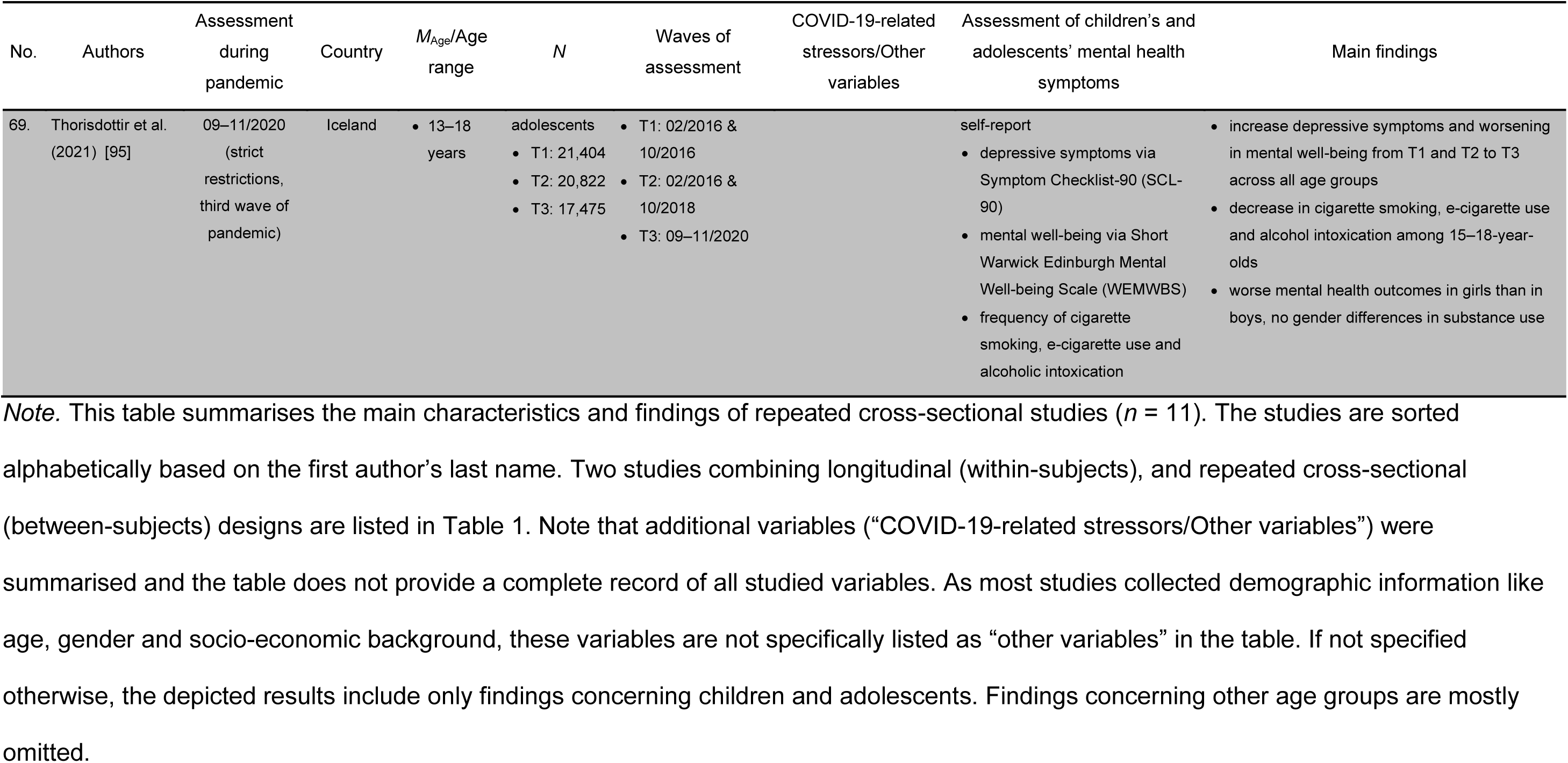
Overview of Repeated Cross-Sectional Studies (Different Samples).

### Changes in mental health due to the COVID-19 pandemic

Table 3 displays the mental health indicators and symptoms assessed in the studies with a pre-pandemic measurement and how frequently the respective indicators and symptoms were assessed. The studies’ findings concerning the change of the prevalence or intensity of the respective mental health indicators and symptoms from before the pandemic to during the pandemic are summarised in Table 3 as well. The seven studies without a pre-pandemic measure are not included in the table [30,33–35,37–39].

**Table 3.** Summary of Main Results of Studies Comparing Pre-Pandemic Levels of Mental Health with Measures During the Pandemic.

| Indicator of mental health | Number of studies (N) | Increase from pre-pandemic to pandemic | No change from pre-pandemic to pandemic | Decrease from pre-pandemic to pandemic |
| --- | --- | --- | --- | --- |
| Quality of life/Well-being | 11 | 2<br>[57,89] | 2<br>[87,92] | 8<br>[28,44,50,57,68,69,85,95] |
| Life satisfaction | 3 | 0 | 1<br>[74] | 2<br>[76,80] |
| Resilience | 1 | 0 | 0 | 1<br>[65] |
| Positive affect | 4 | 0 | 1<br>[54] | 3<br>[73,74,76] |
| **Total positive mental health indicators** |  | **2** | **4** | **14** |
| Global measures of mental health problems | 8 | 5<br>[47,55,77,78,81] | 3<br>[47,64,87] | 1<br>[71] |
| Stress | 5 | 4<br>[31,49,50,70] | 0 | 1<br>[91] |
| Negative affect | 4 | 3<br>[54,73,74] | 1<br>[76] | 0 |
| Internalising symptoms | 21 | 8<br>[36,52,56,58,62,63,78,88] | 9<br>[40,41,45,46,64,77,80,81,92] | 5<br>[55,62,66,71,89] |
| Externalising symptoms | 13 | 8<br>[36,56,58,63,77,78,83,88] | 4<br>[40,41,71,92] | 2<br>[71,89] |
| Depression symptoms | 29 | 19<br>[28,29,42–44,46,51,53,59–61,67,73,74,76,83,84,94,95] | 7<br>[31,58,79,82,86,92,93] | 3<br>[45,75,91] |
| Anxiety symptoms | 20 | 13<br>[28,29,44,51,53,60,72,73,76,79,84,93,94] | 8<br>[31,32,46,51,72,79,83,93] | 5<br>[31,32,45,61,75] |
| Cognitive symptoms | 7 | 5<br>[29,52,58,66,78] | 2<br>[55,71] | 0 |
| Hyperactivity symptoms | 8 | 5<br>[52,58,62,64,77] | 3<br>[29,55,58] | 1<br>[62] |
| Disruptive behaviour symptoms | 9 | 4<br>[29,55,58,62] | 6<br>[29,52,58,64,77,82] | 2<br>[62,92] |
| Behavioural addiction symptoms (electronic media, Internet) | 5 | 3<br>[31,79,90] | 1<br>[79] | 1<br>[49] |
| Substance use and addiction symptoms | 3 | 1<br>[27] | 1<br>[48] | 3<br>[27,48,95] |
| Psychosomatic symptoms | 4 | 3<br>[28,50,66] | 0 | 1<br>[80] |
| Post-traumatic stress symptoms | 2 | 1<br>[83] | 1<br>[93] | 0 |
| Suicidal symptoms | 2 | 0 | 1<br>[92] | 2<br>[91,92] |
| Neurotic symptoms | 1 | 1<br>[42] | 0 | 0 |
| Dissociative symptoms | 1 | 0 | 1<br>[42] | 0 |
| Eating disorder symptoms | 1 | 0 | 1<br>[92] | 0 |
| Psychotic symptoms | 1 | 1<br>[84] | 0 | 0 |
| **Total negative mental health indicators** |  | **84** | **49** | **27** |

Note. This table depicts the number of studies investigating a respective positive or negative indicator of mental health; the number of studies finding a significant increase, significant decrease, and/or no change of a respective indicator from a pre-pandemic assessment to an assessment during the COVID-19 pandemic; and the sum of findings over all positive/negative indicators. Studies with no pre-pandemic measurement are not included in this table ( $n = 7$ ). Studies appear multiple times in the same category if they reported multiple trends, e.g., for different age groups or for different aspect of a symptom group. If a study assessed multiple outcomes, each finding was listed separately.

The results of the 14 studies investigating changes over the course pandemic are summarised hereinafter. Even though the results are mixed, the overall tendency of findings indicates a decrease of mental health symptoms (internalising and externalising symptoms, stress, depression, anxiety, conduct problems, attention problems, problematic smartphone and Internet use) and an increase of well-being, when assessments taken during periods of lockdown/stronger restrictions and higher infection rates were compared with assessments taken during or after the lift of restrictions and easing of the pandemic situation [29–31,33– 35,37,38,40]. However, eight studies did not find a significant change in certain symptoms (i.e. internalising and externalising symptoms, depression, anxiety, attention problems, substance use) and thus did not completely support this trend [27,29,31–35,39]. Two of those studies even reported an increase of certain mental health symptoms (i.e. eating disorder symptoms, hyperactivity, conduct problems) [34,35].

One study comparing measurements between a period of lifted regulations and a following period of reinforced restrictions reported mainly an increase in symptoms of poor mental health (i.e., anxiety, depression, and psychosomatic symptoms, decrease of health-related quality of life) [28].

Although most studies reported a decrease in at least some psychopathological symptoms after an ease of the pandemic situation, it is less clear if children and adolescents return to a pre-pandemic level of functioning. Of the seven studies with multiple measurements during the pandemic and a pre-pandemic assessment, five reported an increase of some symptoms from the pre-pandemic measure to the assessment during the pandemic with loosened restrictions [27–29,31,36], three reported a decrease of symptoms from pre-pandemic levels to during less-restricted periods of the pandemic [27,31,32], and three did not find a difference between the two time points [29,31,40].

### Factors influencing COVID-19-related changes in mental health

Table 4 summarises influencing factors that have been reported to be associated with mental health outcomes during the COVID-19 pandemic and therefore mediate or moderate the effect of the COVID-19 pandemic on children’s and adolescents’ mental health. For simplicity purposes, moderators, mediators, and correlates of outcomes were classified as “risk factors” if they were associated with more negative mental health effects and as “protective factors” if they were associated with less severe negative mental health outcomes.

**Table 4.** Risk and Protective Factors of the COVID-19 Pandemic’s Mental Health Effects.

| Factor categories | Risk factors | Protective factors |
| --- | --- | --- |
| Sociodemographic factors | female gender (depression, anxiety, internalising symptoms, well-being, life satisfaction, stress, psychosomatic symptoms) [37,59–62,68,70,80,83,93,95]<br>male gender (general mental health problems, quality of life, attention problems, addictive behaviour) [28,37,47,90]<br>younger age (behavioural and emotional problems, psychosomatic symptoms, quality of life) [28,35,37]<br>older age (depression, anxiety, behavioural addiction) [27,32,58,66,79,93]<br>being an only child [62,72]<br>low family income, financial worries [29,56,62,70]<br>social disadvantage [28,78]<br>living in an apartment, without access to green space [68] | female gender (general mental health problems, quality of life, attention problems, addictive behaviour) [28,37,47,90]<br>male gender (depression, anxiety, internalising symptoms, well-being, life satisfaction, stress, psychosomatic symptoms) [37,59–62,68,70,80,83,93,95]<br>younger age (depression, anxiety, behavioural addiction) [27,32,58,66,79,93]<br>older age (internalising and externalising problems, quality of life) [28,35,37]<br>living in a house, access to green space [68] |
| Intra-individual factors | pre-existing mental health problems [62,76,83]<br>poor physical health [49]<br>neurodevelopmental disorders, special educational needs [35,37]<br>negative coping strategies (emotion-focussed engaged and disengaged coping) [41,63]<br>dysfunctional/missing emotion regulation abilities (e.g., dampening positive emotions) [29,54,70,74]<br>negative cognitive styles [59]<br>emotional instability [42]<br>high extraversion [43] | good physical health [49]<br>positive coping mechanisms (e.g., talking to friends, prioritising sleep, problem-focussed engaged coping) [51]<br>positive emotion regulation abilities, high emotional awareness/competence [29,54,70,75]<br>resilience [59,65]<br>self-efficacy [63]<br>eudemonic and hedonic motives [74] |
| Parental/Family factors | single parent [62,72,94]<br>parental stress and strain [30,33,70]<br>parental mental health problems, psychological distress [28,35]<br>depression [46,78]<br>anxiety [88]<br>use of alcohol and drugs [27]<br>parental negative coping strategies [41]<br>maternal burden of care [46]<br>parental over-reactivity [41]<br>parental expressive suppression of emotions [33]<br>dysfunctional parenting styles [55]<br>family stress and instability [66] | family functioning [71]<br>positive family climate [28]<br>maternal high mind-mindedness [88] |
| Social factors | deterioration of relationships to others [55]<br>missing friends [66] | friend support/contact to friends [45,46,61,65]<br>social support [28,65,76] |
| COVID-19-related factors | infection risk, infected friend or relative [72,94]<br>perceived lifestyle impact of the pandemic [53]<br>negative change in parental working situation [94]<br>perceived stress [41,59,61]<br>isolation and loneliness [42,43]<br>worry, concerns about health [42,55,61,66]<br>concerns/fears about the pandemic [80,86]<br>frustration, boredom [55]<br>negative home-schooling experience [66]<br>increased restrictions [33,34]<br>feelings of uncertainty [27]<br>material hardship [27]<br>high exposure to news [36] |  |
| Behavioural factors | mainly staying at home during confinement [70]<br>increased, passive screen time [36,78]<br>taking part in many leisure time activities prior to the pandemic [77]<br>less school assignment completion [78] | physical health behaviours:<br>exercising for minimum 1h/day [72]<br>adequate sleep [36]<br>structured routines [36,76]<br>time spent in nature [36] |

Table 5 portrays the change of mental health indicators and symptoms from before to during the pandemic and how frequently these symptoms were assessed in three separate age groups (< six years, six–12 years, > 12 years).

**Table 5.** Summary of Main Results of Studies Comparing Pre-Pandemic Levels of Mental Health with Measures During the Pandemic Sorted by Age Group.

| Indicator of mental health | Direction of Change | Younger children (~ < 6 years) | School-aged children (~ 6-12 years) | Adolescents (~ > 12 years) |
| --- | --- | --- | --- | --- |
| Quality of life/well-being | + | 0 | 1 [89] | 1 [57] |
|  | 0 | 0 | 1 [87] | 1 [92] |
|  | – | 1 [96] | 3 [28,68,96] | 8 [28,44,50,57,68,69,95,96] |
| Life satisfaction | + | 0 | 0 | 0 |
|  | 0 | 0 | 0 | 1 [74] |
|  | – | 0 | 0 | 2 [76,80] |
| Resilience | + | 0 | 0 | 0 |
|  | 0 | 0 | 0 | 0 |
|  | – | 0 | 0 | 1 [65] |
| Positive affect | + | 0 | 0 | 0 |
|  | 0 | 0 | 1<br>[54] | 1<br>[54] |
|  | – | 0 | 0 | 3<br>[73,74,76] |
| **Total positive mental health indicators** | **+** | **0** | **1** | **1** |
|  | **0** | **0** | **2** | **3** |
|  | **–** | **1** | **3** | **14** |
| Global measures of mental health problems | + | 2<br>[47,77] | 3<br>[47,78,81] | 2<br>[55,72] |
|  | 0 | 2<br>[47,64] | 2<br>[47,87] | 0 |
|  | – | 0 | 1<br>[71] | 1<br>[71] |
| Stress | + | 0 | 3<br>[31,49,70] | 3<br>[31,50,70] |
|  | 0 | 0 | 0 | 0 |
|  | – | 0 | 0 | 1<br>[91] |
| Negative affect | + | 0 | 1<br>[54] | 3<br>[54,73,74] |
|  | 0 | 0 | 0 | 1<br>[76] |
|  | – | 0 | 0 | 0 |
| Internalising symptoms | + | 1<br>[88] | 5<br>[36,56,58,62,78] | 4<br>[36,52,62,63] |
|  | 0 | 3<br>[58,64,77] | 5<br>[40,41,45,46,81] | 3<br>[40,80,92] |
|  | – | 0 | 4<br>[62,66,71,89] | 3<br>[55,62,71] |
| Externalising symptoms | + | 2<br>[77,88] | 5<br>[36,56,58,78,83] | 2<br>[36,63] |
|  | 0 | 1<br>[58] | 3<br>[40,41,71] | 3<br>[40,71,92] |
|  | – | 0 | 2<br>[71,89] | 1<br>[71] |
| Depression symptoms | + | 0 | 4<br>[28,46,83,94] | 17<br>[28,29,42–44,51,53,59–61,67,73,74,76,84,94,95] |
|  | 0 | 1<br>[58] | 3<br>[31,58,79] | 7<br>[31,51,79,82,86,92,93] |
|  | – | 0 | 2<br>[45,75] | 2<br>[75,90] |
| Anxiety symptoms | + | 0 | 2<br>[28,94] | 13<br>[28,29,44,51,53,60,72,73,76,79,84,93,94] |
|  | 0 | 0 | 5<br>[31,32,46,79,83] | 4<br>[31,51,72,93] |
|  | – | 0 | 4<br>[31,32,45,75] | 3<br>[31,61,75] |
| Cognitive symptoms | + | 0 | 3<br>[58,66,78] | 2<br>[29,52] |
|  | 0 | 0 | 1<br>[71] | 2<br>[55,71] |
|  | – | 0 | 0 | 0 |
| Hyperactivity symptoms | + | 2<br>[64,77] | 2<br>[58,62] | 2<br>[52,62] |
|  | 0 | 1<br>[58] | 0 | 2<br>[29,55] |
|  | – | 0 | 1<br>[62] | 1<br>[62] |
| Disruptive behaviour symptoms | + | 0 | 3<br>[58,62,83] | 3<br>[29,55,62] |
|  | 0 | 3<br>[58,64,77] | 2<br>[58] | 3<br>[29,52,82] |
|  | – | 0 | 1<br>[62] | 2<br>[62,92] |
| Behavioural addiction symptoms (electronic media, Internet) | + | 0 | 2<br>[31,90] | 3<br>[31,79,90] |
|  | 0 | 0 | 1<br>[79] | 0 |
|  | – | 0 | 1<br>[49] | 0 |
| Substance use and addiction symptoms | + | 0 | 1<br>[27] | 1<br>[27] |
|  | 0 | 0 | 0 | 1<br>[48] |
|  | - | 0 | 1<br>[27] | 3<br>[27,48,95] |
| Psychosomatic symptoms | + | 0 | 2<br>[28,66] | 2<br>[28,50] |
|  | 0 | 0 | 0 | 0 |
|  | - | 0 | 0 | 1<br>[80] |
| Post-traumatic stress symptoms | + | 0 | 1<br>[83] | 0 |
|  | 0 | 0 | 0 | 1<br>[93] |
|  | - | 0 | 0 | 0 |
| Suicidal symptoms | + | 0 | 0 | 0 |
|  | 0 | 0 | 0 | 1<br>[92] |
|  | - | 0 | 0 | 2<br>[91,92] |
| Neurotic symptoms | + | 0 | 0 | 1<br>[42] |
|  | 0 | 0 | 0 | 0 |
|  | - | 0 | 0 | 0 |
| Dissociative symptoms | + | 0 | 0 | 0 |
|  | 0 | 0 | 0 | 1<br>[42] |
|  | - | 0 | 0 | 0 |
| Eating disorder symptoms | + | 0 | 0 | 0 |
|  | 0 | 0 | 0 | 1<br>[92] |
|  | - | 0 | 0 | 0 |
| Psychotic symptoms | + | 0 | 0 | 1<br>[84] |
|  | 0 | 0 | 0 | 0 |
|  | - | 0 | 0 | 0 |
| Total negative mental health indicators | + | 7 | 37 | 59 |
|  | 0 | 11 | 22 | 30 |
|  | - | 0 | 17 | 20 |

Note. This table depicts the number the number of studies finding a significant increase (indicated by “+”), significant decrease (indicated by “-”), and/or no change (indicated by “0”) of a respective indicator from a pre-pandemic assessment to an assessment during the COVID-19 pandemic and the sum of findings over all positive/negative indicators sorted by age of participants. Studies with no pre-pandemic measurement are not included in this table ( $n = 7$ ). Studies might appear multiple times in the same category if they reported multiple trends, e.g., for different age groups or for different aspect of a symptom group. If a study assessed multiple outcomes, each finding was listed separately.

Table 6 summarises regional differences in mental health outcomes by listing those countries contributing more than three studies to this review.

**Table 6.**
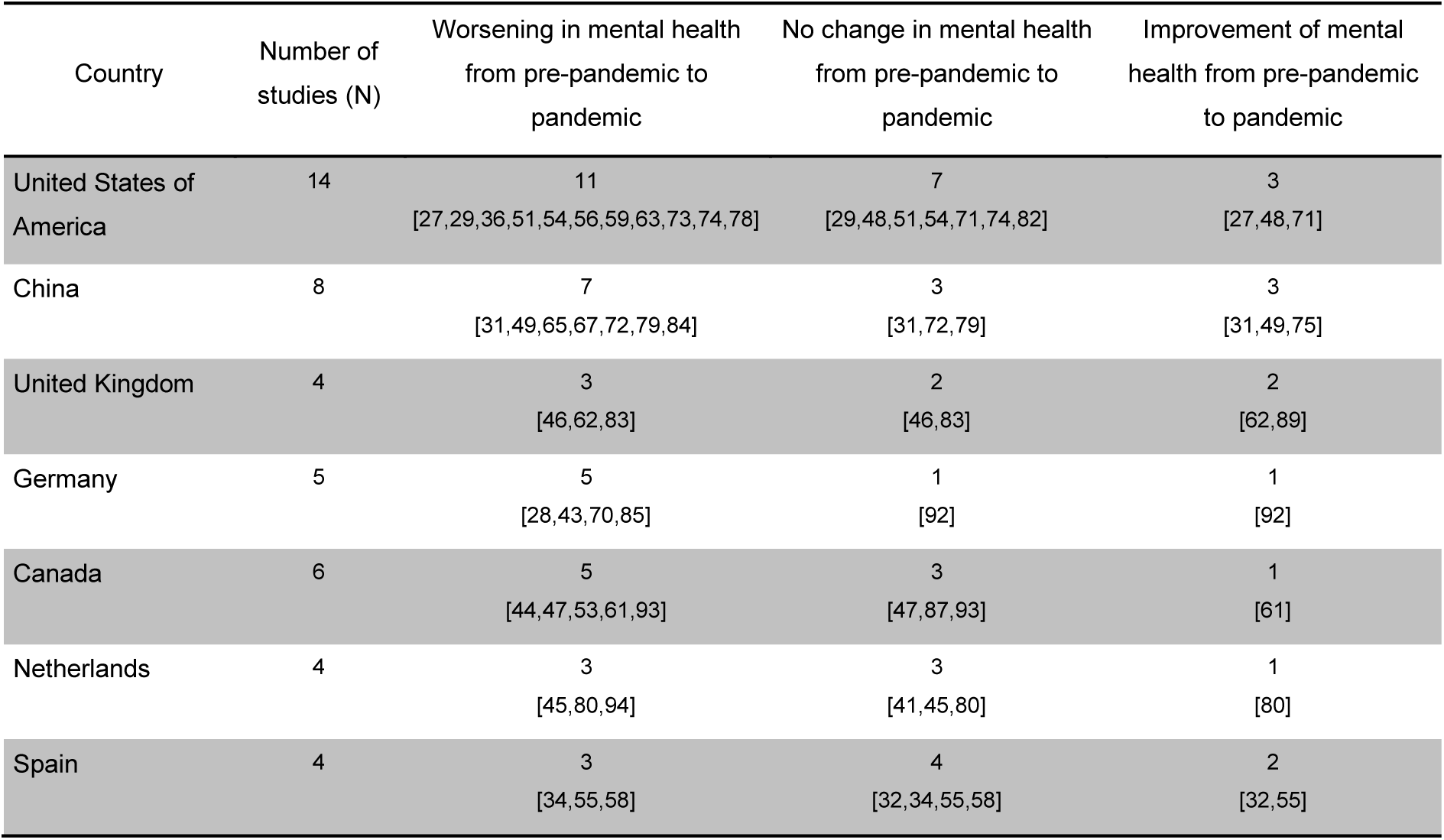
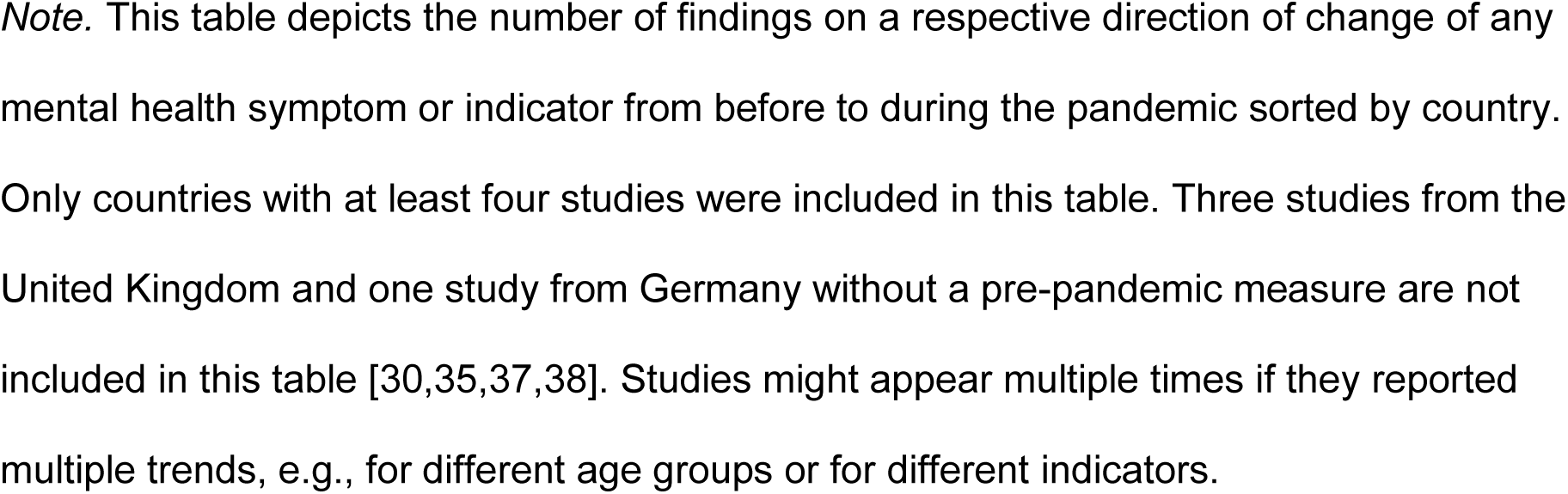
Change of Mental Health from Pre-Pandemic to During the Pandemic Sorted by Country.

| Country | Number of studies (N) | Worsening in mental health from pre-pandemic to pandemic | No change in mental health from pre-pandemic to pandemic | Improvement of mental health from pre-pandemic to pandemic |
| --- | --- | --- | --- | --- |
| United States of America | 14 | 11<br>[27,29,36,51,54,56,59,63,73,74,78] | 7<br>[29,48,51,54,71,74,82] | 3<br>[27,48,71] |
| China | 8 | 7<br>[31,49,65,67,72,79,84] | 3<br>[31,72,79] | 3<br>[31,49,75] |
| United Kingdom | 4 | 3<br>[46,62,83] | 2<br>[46,83] | 2<br>[62,89] |
| Germany | 5 | 5<br>[28,43,70,85] | 1<br>[92] | 1<br>[92] |
| Canada | 6 | 5<br>[44,47,53,61,93] | 3<br>[47,87,93] | 1<br>[61] |
| Netherlands | 4 | 3<br>[45,80,94] | 3<br>[41,45,80] | 1<br>[80] |
| Spain | 4 | 3<br>[34,55,58] | 4<br>[32,34,55,58] | 2<br>[32,55] |

*Note.* This table depicts the number of findings on a respective direction of change of any mental health symptom or indicator from before to during the pandemic sorted by country. Only countries with at least four studies were included in this table. Three studies from the United Kingdom and one study from Germany without a pre-pandemic measure are not included in this table [30,35,37,38]. Studies might appear multiple times if they reported multiple trends, e.g., for different age groups or for different indicators.

## Discussion

The COVID-19 pandemic and the protection measures to contain its spread have massively changed daily lives of billions of children and adolescents worldwide. Here a systematic literature review on the longitudinal effects of the COVID-19 pandemic on child and adolescent mental health was conducted in the face of a lack of reviews and meta-analyses which a) investigate longitudinal changes in child and adolescent mental health, and b) include studies conducted after the initial months of the pandemic. The current review was executed according to the Preferred Reporting Items for Systematic Reviews and Meta-Analyses (PRISMA) statement [26].

Sixty-nine longitudinal and repeated cross-sectional studies published between September 2020 and December 2021 assessing child and adolescent mental health over the first 17 months of the pandemic were included. Changes were assessed in a range of both general mental health indicators and specific psychopathological symptoms from before to during the pandemic and over its course. Furthermore, factors influencing these effects were summarised.

### Changes in mental health from before to during the pandemic

We expected to find an increase in general psychopathological stress levels and symptoms of mental disorders in children and adolescents due to the exposure to the COVID-19 pandemic. This trend was predicted particularly for internalising symptoms of depression and anxiety, externalising symptoms, attentional problems, post-traumatic stress, and psychosomatic symptoms.

Regarding all negative indicators of mental health, such as psychological distress, negative affect, or specific psychopathological symptoms, 84 findings of an increase in a single mental health indicator were reported. However, almost as many findings did not support an increase in negative mental health indicators, with 49 findings showing no change and 27 findings showing a decrease in such an indicator. The findings for the change of global measures of poor mental health (i.e., outcomes not specific to a certain indicator or symptom group, e.g., total scores of mental health questionnaires) are also mixed but more in line with an increase of these constructs, with five findings supportive of this trend compared to four non-supportive findings. There is clearer support for an increase in the indicators “stress” and “negative affect”. Each had only one finding not supportive of an increase, compared to four and three findings supporting an increase, respectively.

The findings for positive mental health indicators, such as quality of life, life satisfaction, positive affect, resilience, and well-being, are more homogeneous and support a decrease of mental health from before to during the pandemic (14 supportive findings vs. six non-supportive findings).

The trend for global measures of internalising symptoms is less clear and would rather suggest no change (nine supportive findings) or an increase (eight supportive findings) than a decrease in symptoms (five supportive findings). This is contradictory to the expectations that predicted a clear increase in internalising symptoms.

Negative effects of the pandemic are clearer if symptoms of individual internalising syndromes are inspected separately. The support for an increase in depressive symptoms is strong (19 supportive findings vs. 10 non-supportive findings). This increase fits the presumed effect based on previous literature and appears logical in the face of enforced isolation, the lack of social contacts, pandemic-related worries, the loss of many positive leisure time activities and daily structure.

However, there is more variability in data regarding anxiety symptoms with 13 studies finding an increase in symptoms and the same number of studies not reporting an increase in symptoms.

A rise in the general scores of externalising symptoms from before to during the COVID-19 pandemic is supported by most of the reviewed studies assessing these symptoms (eight supporting findings vs. six non-supporting findings).

When specific externalising syndromes are regarded separately, different results can be observed. While symptoms of hyperactivity and inattention seem to have increased (hyperactivity: five supportive findings vs. four non-supportive findings; inattention: five supportive vs. two non-supportive findings), there is greater evidence for no change in the frequency of disruptive behaviour symptoms (six supportive findings) than for an increase (four supportive findings) or a decrease (two supportive findings).

The increase in hyperactive symptoms can be explained by the restricted opportunities for physical activity during the pandemic due to stay-at-home orders, closures of schools, sport clubs, playgrounds, gyms, public swimming pools, etc., and the cancellation of leisure time activities associated with physical exercise.

The lack of evidence indicating an increase in disruptive behaviour problems can be explained by the stability of underlying disorders and dispositions. Furthermore, the display of such symptoms is often situation-specific, meaning symptoms only become evident in certain social settings, for example, in school. The exposure to these situations was reduced in the pandemic due to stay-at-home orders and the closures of schools potentially leading to limited chances to exercise and observe these problematic behaviours.

Taken together, in light of the reviewed studies, the first hypothesis of increased psychopathological stress and symptom levels, particularly regarding symptoms of depressive, anxiety, attentional, psychosomatic and post-traumatic stress symptoms, was supported.

The findings are in line with previous research finding an increase in psychopathological distress and symptoms of depression and anxiety in children, adolescents, and adults in the context of disasters in general [97–100] and the COVID-19 pandemic in particular [6,15,16,20–22,101,102].

However, the few existing meta-analytic studies regarding mental health effects of the COVID-19 pandemic based on longitudinal data [23–25] indicated no or only slight changes in mental health in the general population due to highly heterogeneous data. This high variability in findings across studies was also observed in this review, in particular regarding more global measures of mental health indicators (e.g., internalising symptoms). Negative effects of the pandemic on children’s mental health became more detectable when specific symptom groups were examined separately and when studies were sorted by age of participants. This fits the assumption that effects might differ across different regions, social groups, and contexts which will be discussed below.

### Changes in mental health due to changes in pandemic intensity and restrictions

We expected that pandemic-related health protection measures, in particular confinement and quarantine, should have a negative effect on child and adolescent mental health. Consequently, a reduction in restrictions over the course of the pandemic should be associated with decreasing symptoms of poor mental health.

Not all reviewed studies assessing the changes in mental health indicators and symptoms over the course of the pandemic support this hypothesis. Nonetheless, more findings are in favour of a decrease of symptoms after a lift of restrictions (*n* = 9) than of no change in symptoms (*n* = 8) or an increase in symptoms (*n* = 2).

This conclusion is in line with data from the newest wave of one of the studies reviewed here by Ravens-Sieberer et al. (2022) [103]. This German study shows that levels of low health-related quality of life and internalising symptoms in autumn 2021 were still higher than pre-pandemic levels. However, health-related quality of life and mental health improved from spring 2020 and winter 2020/21 to autumn 2021. The authors explain this effect by lower infection rates, higher vaccination rates, and loosening of restrictions.

Reasons for the negative effect of quarantine, confinement, and lockdown on child and adolescent mental health are diverse. For example, Mohler-Kuo et al. (2021) [104] found in their survey on stress factors during lockdown in Switzerland that the most common sources for stress during lockdown were the disruption of social life, the break-down of normal routines, the cancellation of important plans and events, and the uncertainty and unpredictability of the duration of the pandemic aside from distressing news coverage and rapidly changing recommendations as well as the fear of infection and the pandemic itself. Overall, it must be considered, that protection measures usually correlate highly with pandemic intensity and infection rates. Contrary, restrictions are usually lifted when infection rates ease. Thus, a unique contribution of the lifting of containment measures independent from infection rates can not be assumed [105].

### Changes of mental health related to individual risk and protective factors

Overall, the included studies suggest low socio-economic status, financial worries, material hardship, lack of space, negative home-schooling experience, bad physical health, and the diagnosis of a neurodevelopmental disorder as key risk factors for experiencing stronger negative mental health effects due to the pandemic (see Table 5 for detailed references).

There was also evidence that some children and adolescents that were less likely than their healthy peers to display an increase in psychopathological distress and symptom levels in response to the onset of the pandemic. These children and adolescents are reported to have experienced constant high levels of psychological distress, before and during the pandemic, due to early childhood stress, maltreatment experiences, certain chronic mental health problems, special needs, and socio-economic disadvantage [37,51,60,62]. Their psychological distress was also mostly unrelated with the changing infection rates and health protection measures. This means that they did not profit from the lifting of restrictions or the easing of the pandemic situation as their same-aged peers did. This is a rather unexpected finding in the light of previous research, as some of these factors, such as prior traumatic experiences or dependency on special psychological support, have been previously proposed to enhance the risk of experiencing negative mental health effects in the face of a disaster [22,100,106].

We further investigated age effects by grouping the reviewed studies by age of the participants (see Table 4 for detailed references). Most evidence for an increase in mental health problems (59 supportive vs. 50 non-supportive findings) and decrease of positive mental health indicators (14 supportive findings vs. four non-supportive findings) was reported for the age group of adolescents (i.e., individuals approximately 12 to 18 years of age). In this age group strong and convincing support for an increase in depressive (17 supportive findings vs. nine non-supportive findings) and anxiety symptoms (13 supportive vs. seven non-supportive findings), and a decrease in quality of life and well-being (eight supportive findings vs. two non-supportive findings) was found.

A general trend of increasing psychopathological distress and symptoms could also be found in school-aged children (i.e., individuals approximately six to 12 years of age) with 37 studies reporting an increase in mental health problems, 22 not finding a change in symptoms, and 17 reporting a decrease in negative mental health indicators. In contrast to adolescents, no clear evidence for an increase in depressive and anxiety symptoms were found in this younger age group.

Hence, adolescents might have suffered more strongly from reduced contact to peers and from heightened demands for personal responsibility (e.g., self-directed learning).

Additionally, literature on the psychological development in childhood and adolescence has stated that adolescents are more vulnerable to social stressors, such as isolation and loneliness [107] and that symptoms of affective disorders rise significantly in adolescence [108,109]. This age-related difference might also be partially due to the increasing introspective ability of adolescents that allows them to report more reliably on internalising symptoms than children. Another reason might be that parent-reports, typically associated with an underreporting of internalising symptoms, were used for children but not for adolescents.

Concerning younger children (i.e., individuals approximately under six years of age), most studies does not indicate a change in psychopathological distress and symptoms (11 supportive vs. eight non-supportive findings). However, this age group is severely under-investigated, and the reliability and validity of results might be limited due to difficulties in the assessment of symptoms in such young children and the mere reliance on parent- or educator-reports in this review. Therefore, it is not possible to draw clear conclusions for this age group, although it seems that it is less affected by negative mental health effects of the pandemic than older age groups.

Gender effects were not consistently investigated and reported in the reviewed studies. Of the 14 studies finding and reporting a significant gender effect, 11 studies identified female gender as a risk factor for higher levels and/or a stronger increase in certain indicators of poor mental health and five studies identified male gender as such a risk factor (see Table 5 for detailed references).

Females were reported to be at a higher risk for higher levels and/or stronger increases in internalising, anxiety, and depressive symptoms, stress, and lower levels of well-being than males. At the same time, males seem to have been more prone to attention problems, addictive gameplay, and sharper decreases in quality of life and life satisfaction than females. These gender differences were nearly exclusively found in adolescent samples.

Regarding socio-economic variables, children and adolescents who grow up with a single parent and those who do not have siblings were particularly at risk to show decreased levels of mental health during the pandemic. Further, housing situations providing only limited living space and no access to green spaces were associated with higher increases in and/or levels of mental distress. A low socio-economic status and financial worries of the family were also identified as an important risk factor, especially if the children did not already experience heightened psychological distress prior to the pandemic.

Other individual factors that acted as risk factors for mental health include poor physical health prior to the pandemic, the diagnosis of a neurodevelopmental disorder, and dysfunctional or lacking coping and emotion regulation strategies.

Parental strain and psychological distress, particularly in the form of parental symptoms of anxiety, depression, and substance abuse were found to be an important influential factor for child and adolescent mental health during the pandemic. Further risk factors for poor mental health in the family environment include negative parental coping strategies, dysfunctional parenting styles, and overall family stress and instability.

Pandemic-specific factors leading to more psychological distress were the perceived lifestyle impact of the pandemic-related policies on the one hand, and stress due to the pandemic situation on the other. The first includes negative changes in the parental job situation, the experience of isolation and loneliness, frustration and boredom, and negative experiences during home-schooling. The second comprises fears of infection with the virus concerning oneself or family and friends, general health-related concerns, and worries due to the uncertainty of the pandemic situation.

Behavioural factors that contributed to negative mental health outcomes were reported to be prolonged screen time, less physical exercise, disrupted sleep patterns, and mainly staying inside during times of confinement.

Factors that promote mental health or to mitigate negative mental health effects include access to green spaces and time spent in nature during times of confinement. Good physical health and health-related behaviours, such as regular physical exercise, a healthy diet, structured routines, and regular sleep patterns were reported to help to protect child and adolescent mental health. Children’s and adolescents’ positive and adaptive coping mechanisms and emotion regulation abilities as well as social support through families and friends have also been identified as protective factors. Family functioning and a positive family climate have been found to help reduce negative mental health effects.

### Reasons for inconsistencies in findings

The variability in the study sample, for example, considering times and locations (i.e., country/region) of assessment, might be a strong factor for the inconsistency of findings among the reviewed studies. Differing phases of the pandemic and locations of assessment are related to variations in infection rates and health protection measures as well as to the duration and intensity of exposure to the pandemic at the time of assessment.

The timing of assessments is particularly important in studies investigating changes in symptoms over the course of the pandemic. An explanation for the fact that some studies could not find a decrease in symptoms after a lift of restrictions might be that recovery takes time and therefore might not be immediately visible in the assessments. Some stressors named above, such as unpredictability of the situation and fear of infection, have most likely continued to be a burden to mental health even with lifted restrictions.

Further influencing factors leading to higher heterogeneity in findings might have been differences in sample characteristics (e.g., age, socio-economic status), methodological approaches, the conceptualisation of mental health problems, and the definition and operationalisation of outcome variables.

Another important reason for the heterogeneous findings on the effects of health protection measures might be that mental health is not only influenced by the severity of protection measures but also associated with other characteristics of the pandemic (e.g., infection rates, death tolls). It is difficult to distinguish the effects of the intensity of protection measures from those of the severity of infection rates and the perceived pandemic threat. For example, one of the reviewed studies comparing mental health effects between three European countries showed a higher increase of mental health problems in those countries with stricter restrictions, which, however, also were the countries with higher infection and death rates at the time [34].

In fact, it has been proven that both policy stringency and pandemic intensity affect mental health to a similar degree [105]. This means that minimising transmission of the virus and death rates by potentially stricter health protection measures might indeed be protective against negative mental health effects due to the pandemic’s intensity.

It seems more appropriate to use separate measures for distinct symptom categories (e.g., depressive symptoms, psychosomatic symptoms) than global measures (e.g., internalising symptoms), as the latter might be less suited to accurately capture a change in single symptoms. The same explanation can be used to explain the heterogeneous findings for complex symptom groups (e.g., anxiety) that comprise many different forms of a syndrome (e.g., generalised anxiety, health-related anxiety, school-related anxiety) that might have developed differently during the pandemic (see e.g., 93).

Furthermore, there is evidence for the existence of age-related differences in the change of mental health symptoms due to the pandemic. For instance, it seems that adolescents have experienced an increase in anxiety symptoms while children have not. Summing up these different effects in the total study sample might have contributed to the heterogeneity of findings.

### Strengths and limitations of the current review

To our knowledge our review is the first which only includes studies with multiple assessment waves, which allows for the detection of changes in mental health which can be clearly attributed to the COVID-19-pandemic. This is a huge advantage in comparison to previous papers which mainly relied on cross-sectional estimations of the prevalence of certain mental health indicators.

Furthermore, this review does not only include studies conducted in the early months but comprises research assessing mental health over the first one and a half years of the pandemic. Therefore, conclusions drawn from this work are not restricted to the initial outbreak and beginning of the pandemic but are suited to estimate more long-term effects.

Additionally, the large study sample size and the inclusion of data from 21 countries in four continents allows for a higher generalisability of results.

There is potential for errors in the study selection as the identified studies were assessed for eligibility, selected, and synthesised by only one researcher. Another reason for possible distortion in data might have been effects of the publication bias due to, for example, the under-publication of non-significant results (“file drawer problem”, see e.g., 110,111). This review only included published and peer-reviewed articles and did not attempt to search for unpublished articles to ensure the scientific quality of studies. However, there is a chance that the results reviewed here therefore represent a biased sample of relevant data.

The conclusions drawn in this paper are most representative of western, educated, industrialised, rich, and democratic societies (WEIRD-bias, see e.g., 112) as most studies were conducted in Northern American and Western European countries. In the face of a global pandemic which has most likely affected socially and economically disadvantaged groups the most, this is a serious limitation to this review. ost of the studies in the sample concentrated on the first months of the pandemic, i.e., March to June 2020.

Furthermore, two-thirds of all included studies were conducted during stricter confinement rules. Due to this lack of studies conducted after summer 2020 and outside heightened restrictions, long-term effects of the pandemic situation, which lasted way longer after this first initial period, are not completely evaluable.

The sample is most representative of early adolescent years and does not allow for clear statements about effects on younger age groups, especially about early childhood.Lastly, the large heterogeneity of time and location of assessment, of current pandemic severity and protection measures at the time of assessment, of methodological approaches (e.g., over 60 different assessment tools were used), and of studied age groups, outcomes, and covariates makes it difficult to draw coherent conclusions.

A major limitation of this literature review is that all included studies rely only on assessments of mental health via child or adolescent self-, caretaker-, or teacher-reports on questionnaires which were presented online in most cases. None of the included studies used clinical evaluations by mental health professionals (e.g., diagnostic interviews), which would be necessary to form solid clinical diagnoses [113]. Consequently, the trends observed in this study do not allow for conclusions about the change of the prevalence of diagnoses but are rather suited to estimate changes in the frequency of mostly self- or parent-reported symptoms. Moreover, the lack of studies using multiple informants to report on a child’s or adolescent’s mental health limits the validity of results.

The methodological quality of the reviewed studies was neither assessed nor accounted for. Effect sizes of findings were not analysed. This reduces the options for summarising the findings to the qualitative method of vote-counting which is substantially inferior to quantitative statistical methods of synthesising research findings. Hence, the purpose of this paper should be seen in an accumulation and summary of the research currently available and an indication of general directions of changes in child and adolescent mental health due to the COVID-19 pandemic.

### Implications for intervention strategies to meet mental health needs of children and adolescents

This paper shows that children’s and adolescents’ psychological distress and symptoms of mental health disorders increased from before to during the pandemic. Missed chances and milestones of development and the delay in diagnosis and treatment will have long-term mental health consequences. Consequently, it is obvious that the need for psychological care for children and adolescents has risen and is still rising worldwide. The situation of mental health care has already been precarious before the pandemic. It has now become even more painfully evident that there is a great dissonance between the demand for and the availability of mental health care in many places worldwide. Therefore, it is important to invest in research into, creation, and implementation of intervention strategies to react to this growing need of psychological care.

Furthermore, this review has shown that it is important to find a balance between health protection and infection control measures on the one side, and the protection and guarantee of the societal participation of children and adolescents on the other.

In general, continuity and stability of access to school, mental and physical health care, and social services are essential for children’s and adolescents’ mental health. Further disruptions should be avoided at all costs. Decisions about the implementation of certain interventions should always be based on expertise and empirical research to allow for efficiency in both the prevention of infections and protection of children’s and adolescents’ mental well-being

### Implications for further research

There is a continued need for preferably longitudinal studies examining the long-term effects of the prolonged pandemic situation on child and adolescent mental health, as studies investigating mental health effects on the pandemic in its later stages are lacking.

It has become evident in the current paper that there is need for more longitudinal or repeated cross-sectional studies investigating the changes in certain symptom categories, as there were not enough studies identified to draw cohesive conclusions about the development of these symptoms. These categories include psychosomatic, post-traumatic stress, and compulsive symptoms, symptoms of substance abuse and addiction as well as symptoms of behavioural addiction to electronic media.

The current paper focusses on the COVID-19-pandemic’s mental health effects on community samples of children and adolescents. While this helps the generalisability of findings, it does not allow conclusions about the pandemic’s effects on special vulnerable groups. This includes children belonging to marginalised ethnic, religious, or social groups, children of frontline workers, children and adolescents growing up in low-income families, and children and adolescents suffering from chronic physical or mental health conditions, such as neurodevelopmental disorders. There is evidence that these groups of children and adolescents have been more strongly affected by the pandemic’s effects than the general population [114–117].

Lastly, it is important to point out that further research into the COVID-19 pandemic’s mental health effects and into strategies for the mitigation of negative outcomes is important far beyond the current health crisis. There is a causal link between climate change and an increase in the frequency of pandemic outbreaks (e.g., 118,119). This means that if we fail to appropriately react and fight against the current climate crisis, we will have to prepare ourselves for future pandemics. Therefore, it is important and urgent to analyse the effects of the current pandemic in depth and to plan and prepare future actions accordingly based on facts and empirical evidence.

## Conclusion

This systematic review of 69 longitudinal and repeated cross-sectional studies has demonstrated that psychopathological distress and symptoms in children and adolescents have increased globally in the first 17 months of the COVID-19 pandemic compared to pre-pandemic data.

The psychological distress of children and adolescents has been shown to be positively associated with both the intensity of the pandemic situation (e.g., infection rates, death tolls) and the severity of protection measures, such as confinement and quarantine.

Among many other factors, female gender, adolescent age, socio-economic disadvantage, parental psychopathology, dysfunctional family environment, social isolation and loneliness, loss of routines and structure, and the experience of distressing emotions due to health-related worries and uncertainty of the pandemic situation have been identified to increase the chances to suffer negative mental health consequences.

Future research is needed to extend the current findings with assessments of the long-term effects of pandemic exposure and of the changes in clinical diagnoses. This is necessary to estimate the increased need for child and adolescent mental health care more concretely. The protection of children’s and adolescents’ mental health must be prioritised at all costs in a crisis like the COVID-19 pandemic to prevent life-long harm to future generations.

## Other Information

This paper was not registered and no protocol was prepared. The authors received no specific funding for this work. The authors have declared that no competing interests exist. All relevant data are within the manuscript and its supporting information files.

## Supporting information

PRISMA Checklist

## Data Availability

All data produced in the present work are contained in the manuscript.

## Supporting information

**S1 Table. Number of Studies with Data Collection Conducted in a Respective Month in the COVID-19 Pandemic.**

**S2 Table. Number of Studies Including Participants in a Respective Age Group.**

**S3 Table. Number of Studies Conducted in a Respective Country.**

