## Supplemental Table 1 for "Systematic review: Longitudinal effects of the COVID-19 pandemic on child and adolescent mental health"

**S1 Table. Number of Studies with Data Collection Conducted in a Respective Month in the COVID-19 Pandemic.**

| Months | Feb 20 | Mar 20 | Apr 20 | May 20 | Jun 20 | Jul 20 | Aug 20 | Sep 20 | Oct 20 | Nov 20 | Dec 20 | Jan 20 | Feb 20 | Mar 20 | Apr 20 | May 20 | Jun 20 |
| --- | --- | --- | --- | --- | --- | --- | --- | --- | --- | --- | --- | --- | --- | --- | --- | --- | --- |
| Number of studies | 1 | 16 | 33 | 37 | 29 | 18 | 11 | 8 | 11 | 13 | 11 | 7 | 4 | 2 | 1 | 1 | 1 |
