## Supplemental Table 2 for "Systematic review: Longitudinal effects of the COVID-19 pandemic on child and adolescent mental health"

**S2 Table. Number of Studies Including Participants in a Respective Age Group.**

| Age in years | 3 | 4 | 5 | 6 | 7 | 8 | 9 | 10 | 11 | 12 | 13 | 14 | 15 | 16 | 17 | 18 |
| --- | --- | --- | --- | --- | --- | --- | --- | --- | --- | --- | --- | --- | --- | --- | --- | --- |
| Number of studies | 7 | 11 | 12 | 13 | 18 | 19 | 25 | 34 | 36 | 43 | 44 | 43 | 39 | 34 | 21 | 10 |

*Note.* Due to missing information about included age ranges in some studies, the numbers presented in this table might not be fully accurate. In case an age range was not indicated, the two age categories closest to the mean age reported in the study were included in this table (e.g., if *M* = 4.6 years, the study was included in the age categories 4 and 5). If no information about age, but about school years was given in a study, this information was used as an estimate of the included age range.
