## Supplemental Table 3 for "Systematic review: Longitudinal effects of the COVID-19 pandemic on child and adolescent mental health"

**S3 Table. Number of Studies Conducted in a Respective Country.**

| Country | USA | China | | United Kingdom | | Germany | | Canada | | Netherlands | | Spain | | Italy | | Norway | | Sweden | | Japan |
| --- | --- | --- | --- | --- | --- | --- | --- | --- | --- | --- | --- | --- | --- | --- | --- | --- | --- | --- | --- | --- |
| Number of studies | 14 | 8 | | 7 | | 6 | | 6 | | 4 | | 4 | | 3 | | 3 | | 2 | | 2 |
| Country | South Korea | | Israel | | Denmark | | Switzerland | | Lithuania | | Ukraine | | Portugal | | Iceland | | Ireland | | Brazil | |
| Number of studies | 2 | | 2 | | 1 | | 1 | | 1 | | 1 | | 1 | | 1 | | 1 | | 1 | |

*Note*. One study was conducted in three countries and thus appears three times in this table Orgilés et al. (2021) [34].
